# Association between pre-existing conditions and hospitalization, intensive care services and mortality from COVID-19 – a cross sectional analysis of an international global health data repository

**DOI:** 10.1101/2023.08.31.23294901

**Authors:** Basant M. S. Elsayed, Lina Altarawneh, Habib Hassan Farooqui, Muhammed Naseem Khan, Giridhara Rathnaiah Babu, Suhail A.R. Doi, Tawanda Chivese

**Affiliations:** Department of Population Medicine, College of Medicine, QU Health, Qatar University, Doha, Qatar

**Author notes:** These authors contributed equally (Basant M. S. Elsayed, Lina Altarawneh). Corresponding author: Tawanda Chivese, Department of population medicine, college of medicine, QU Health, Qatar University, P.O. Box 2713, Doha, Qatar.

**Keywords:** COVID-19 severity, pre-existing conditions, comorbidities, global health, children

## Abstract

**Objective:** To investigate the association between pre-existing conditions and hospitalization, need for intensive care services (ICU) and mortality due to COVID-19.

**Methods:** We used data on all cases recorded in the Global Health Data repository up to the 10^th^ of March 2021 to carry out a cross-sectional analysis of associations between cardiovascular diseases (CVD), hypertension, diabetes, obesity, lung diseases and kidney disease and hospitalization, ICU admission and mortality due to COVID-19. The Global Health repository reported data from 137 countries, but only Brazil, Mexico and Cuba reported more than 10 COVID-19 cases in participants with preexisting conditions. We used multivariable logistic regression to compute adjusted odds ratios (aOR) of the three outcomes for each pre-existing condition in ten-year age groups from 0-9 years and up to 110-120 years.

**Results:** The Global Health repository held 25 900 000 records of confirmed cases of COVID-19, of which 2 900 000 cases were from Brazil, Mexico and Cuba. The overall adjusted odds of hospitalization for the selected pre-existing condition were; CVD (OR 1.7, 95%CI 1.7-1.7), hypertension (OR 1.5, 95%CI 1.4-1.5), diabetes (OR 2.2, 95%CI 2.1-2.2), obesity (OR 1.7, 95%CI 1.6-1.7), kidney disease (OR 5.5, 95%CI 5.2-5.7) and lung disease (OR 1.9, 95%CI 1.8-1.9). The overall adjusted odds of ICU admission for each pre-existing condition were; CVD (OR 2.1, 95%CI 1.8-2.4), hypertension (OR 1.3, 95%CI 1.2-1.4), diabetes (OR 1.7, 95%CI 1.5-1.8), obesity (OR 2.2, 95%%CI 2.1-2.4), kidney disease (OR 1.4, 95%CI 1.2-1.7) and lung disease (OR 1.1, 95%CI 0.9-1.3). The overall adjusted odds of mortality for each pre-existing condition were; CVD (OR 1.7, 95%CI 1.6-1.7), hypertension (OR 1.3, 95%CI 1.3-1.4), diabetes (OR 2.0, 95%CI 1.9-2.0), obesity (OR 1.9, 95%CI 1.8-2.0), kidney disease (OR 2.7, 95%CI 2.6-2.9) and lung disease (OR 1.6, 95%CI 1.5-1.7). The odds of each outcome were considerably larger in children and young adults with these preexisting conditions than for adults, especially for kidney disease, CVD, and diabetes.

**Conclusion:** This analysis of a global health repository confirms associations between pre-existing diseases and clinical outcomes of COVID-19. The odds of these outcomes are especially elevated in children and young adults with these preexisting conditions.

## Background

COVID-19 continues to cause severe illness and deaths, with more than 6.9 million deaths as of 14^th^ of June 2023(1). Although the majority of the people with COVID-19 are likely to have a mild disease course, the absolute numbers of people with severe illness requiring hospitalization, critical care and dying from COVID-19 remains high, driven by the extremely high numbers of people with COVID-19 at a time. This has resulted in pressure on healthcare services, especially hospital and critical care occupancy, causing collateral damage as individuals with other conditions, beside COVID-19, have been deprived of timely and quality care. The availability of efficacious vaccines has reduced the pressure on health services in some high-income countries but low– and middle-income countries (LMICs) have struggled to access vaccines and as a result remain at high risk of repeated surges in COVID-19 infection waves. Within these populations, both children and adults with pre-existing conditions appear to be at risk of worse outcomes from COVID-19.

Studies (2, 3), mostly from high income countries have shown that pre-existing conditions, such as diabetes (4–9), cardiovascular diseases (CVD) (7–10), kidney diseases (8–10), cancer (9) have been consistently associated with poor outcomes from COVID-19. However, findings on the associations between other preexisting conditions such as hypertension and lung diseases are conflicting, with some studies (6–9) showing worse outcomes while others (11) have not found differences in outcomes between people with these conditions and those without.

The Covid-19 pandemic has shown the importance of accurate real-time individual level data, to enable response to the pandemic at different times. In the fight against the pandemic, this builds a stronger understanding of how the virus is spreading, when and how, enabling anticipation of healthcare services use and allocation of health resources. Global data repositories provide anonymized individual level comprehensive list of cases, a platform for sharing information to curb the pandemic and for global risk communication. Global sharing of data likely influence the sharing of best practices in research through means that are reproducible and feedback from critical appraisal of these data systems.

Although a wealth of literature is available on the association between preexisting disease conditions and poor outcomes from COVID-19 infection, there are several gaps in knowledge that still need to be explored. While age is generally accepted to be the strongest risk factor for poor outcomes from COVID-19 (12–14), data on the effect of preexisting condition on outcomes from COVID-19 in different age groups are scarce.

The risk of adverse outcomes of COVID-19 population in children and young adults with comorbidities is not well reported in the literature. A meta-analysis of 42 studies with 9353 children with comorbidities showed that children with comorbidities have a higher risk of COVID-19 and a 3 times higher risk of death from COVID-19, compared to children without comorbidities (13). In this meta-analysis, comorbidities were not analysed separately. Notably, many of the existing primary studies have very small sample sizes, with the exception of two large studies, one from the USA (15) and another from Italy(16). There is also an underrepresentation of data from low and middle income countries (LMICs).

The scarcity of data from LMICs, where other socio-economic determinants of poor outcomes from COVID-19, such as access to screening and treatment for the preexisting are less optimal. In this study, we used an international data repository of cases with COVID-19 from 3 LMICs to investigate the association between preexisting conditions and the following outcomes of COVID-19, 1) hospitalization, 2) need for intensive care services, and 3) mortality.

## Methods

### Study design and setting

In this cross-sectional study, we used data from cases collected in the global health (GH) data repository (17, 18) (https://data.covid-19.global.health/cases). The GH is a collaborative network of volunteers and organizations whose aim is to enable real-time sharing of verifiable data on COVID-19 (18). The full details of the methodology of the GH are described elsewhere(17). In brief, the GH database is an open-source repository which was created to provide reliable real time data of verifiable and anonymized individual level data on confirmed cases with COVID-19 (17). Data from cases were collected using official government reports, such as press releases, official websites, official social media accounts and primary data from peer-reviewed publications. Additional data of individual cases were also collected from other sources such as news websites, news aggregators and peer reviewed publications (17) and used to update data of cases. Data were captured by curators who are skilled in English, Mandarin Chinese, Cantonese, Spanish and Portuguese and machine learning and manual checking was used to reduce duplicate entries and other errors in data capturing (17).

Although a standard data collection template was used in the GH repository, data from only a few countries include preexisting conditions. In this analysis, we included all cases in the GH database collected up to the 10^th^ of March of 2021 from countries where preexisting conditions and the outcomes of interest were reported. We excluded cases where data on country of residence, gender and age were missing. We also excluded cases whose age was categorized into very broad age groups, for example, 0–35 years.

### Data extraction and data management

The GH collects data on geographical location, demographic, dates of confirmation of COVID-19, travel history, preexisting conditions and outcomes for each confirmed case. For each case, we extracted data on the following: age and gender, country of residence, preexisting conditions including diabetes, cardiovascular disease, hypertension, asthma, lung disease, and chronic kidney disease, and the three outcomes of interest which were hospitalization, ICU admission and mortality. A *for-loop* was used to extract data on each of the individual preexisting conditions into separate columns from the column of preexisting conditions in the GH dataset. Data on hospitalization and ICU admission were extracted from the relevant columns and data on death was extracted from the “Outcome” column. For each case, additional data preexisting conditions and outcomes were also extracted from the “Notes” variable using a *for-loop. In the for-loop,* we used text words and synonyms for each of the preexisting condition and outcome in each of the languages used by the GH curators (Supplementary text S1). Data were checked using summaries and frequency tables.

### Data analysis

We described categorical data using frequencies and percentages and compared groups using chi square tests. Within each ten-year age group, we computed unadjusted and adjusted odds ratios (OR) and their 95% confidence intervals (95%CI) for each preexisting condition against the three binary outcomes (hospitalization, ICU admission and mortality) using logistic regression. We adjusted for gender and other preexisting conditions if they were confounders of the association under consideration. For the association between CVD and mortality, we adjusted for gender, diabetes, hypertension, and diabetes. For the association between CVD and mortality, we adjusted for gender, country, diabetes, and obesity. For the association between diabetes and mortality, we adjusted for gender, country, CVD, and obesity. For the association between lung diseases and mortality, we adjusted for gender, country, and obesity. For the association between kidney diseases and mortality, we adjusted for gender, country, CVD, diabetes, and obesity. For the association between obesity and mortality, we adjusted for gender, country, CVD, and diabetes. For the association between hypertension and mortality, we adjusted for gender, country, diabetes, and obesity. The complete Stata do-file for the analysis is in Supplementary Doc S2.

### Ethics

The conduct of this study was guided by the ethical principles of the Declaration of Helsinki (19). Ethics approval and informed consent was not required as the study used deidentified data from a publicly available repository.

## Results

### Characteristics of cases in the whole global health repository

A total of 25 774 885 case records from 137 countries were in the GH repository as of the 10^th^ of March 2021 (Fig. 1). The demographic characteristics of these cases, comparing hospitalizations, ICU and mortality, are shown in Supplementary Table 1. About half (47.3%) of the cases were female, 43.3% were male and 9.2% had missing gender data. Males, compared to females, had higher proportions of hospitalization (3.2% vs 2.8%), ICU admission (0.3% vs 0.2%), and mortality (2.1% vs 1.6%). About three quarters (76.0%) of the cases were in 2020, and a quarter were diagnosed in 2021 (24.0%). The year 2020, compared to 2021, had higher rates of hospitalization (2.9% vs 2.2), ICU admission (0.2% vs 0.1%), and mortality (1.9% vs 1.1%). The highest proportion of cases in the repository were diagnosed during the period of October-December 2020 (40.8%) and the lowest during January-March 2020 (1.8%). The countries with the biggest contributions of cases to the repository were the USA (58.1%), Germany (9.5%), Colombia (8.6%), and Brazil (7.3%).

**Fig 1.**
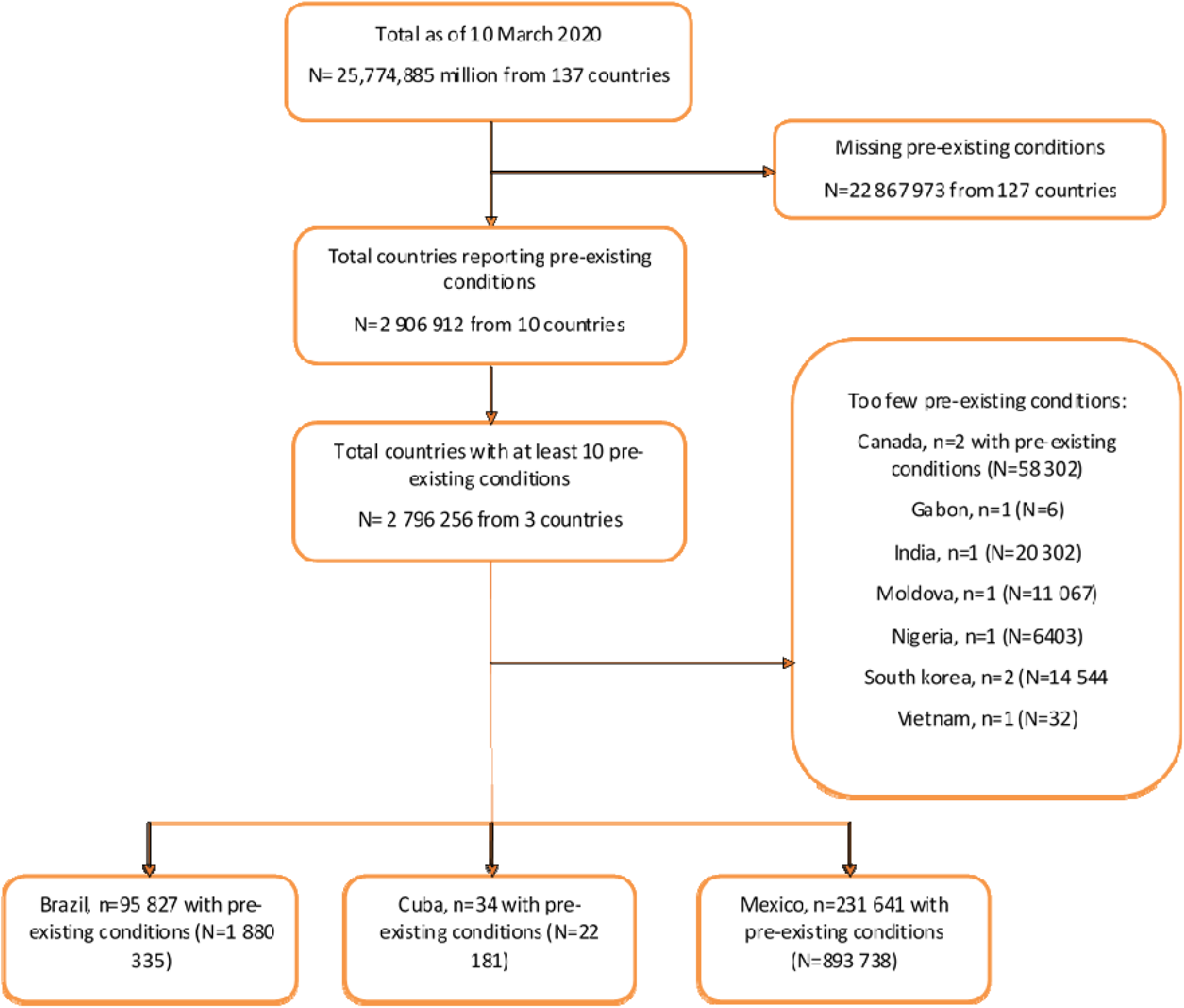
– Flow chart showing the study.

### Characteristics of cases from countries where preexisting conditions were reported

Ten countries had records of pre-existing conditions while the remainder did not have (Fig 1 & Supplementary Table 2). Of these ten countries, only three countries, with a total of 1,919,628 cases, provided data on a substantial number of cases with pre-existing conditions, Mexico (n= 893,167), Brazil (n= 1,015,975) and Cuba (n= 10,486) (Supplementary Table 3), therefore only cases from these countries were considered for the analysis. Out of these cases, 338 cases had missing data on gender and were not included in the analysis, resulting in a total of 1,919,290 (Supplementary Table 3). Just over half of the cases from the countries with pre-existing conditions were from Brazil (52.9%). The characteristics and pre-existing conditions of cases from these three countries are compared by outcome in Supplementary Table 3. Overall, 52.6% were female and 47.4% were male. Compared to females, males had higher proportions of hospitalization (5.1% vs 3.6%, respectively), ICU (0.3% vs, 0.2%, respectively), and mortality (1.8% vs 1.1%, respectively). Most of the cases were aged 30-39 years (22.9%), followed by those aged 40-49 years (19.8%), and 20-29 years (19.4%). The percentage of cases with severe outcomes (hospitalization, ICU ad mortality) increased with increasing age (Supplementary Table 3).

### Comparison of hospitalisations, ICU and mortality by pre-existing condition status

Overall, for all pre-existing conditions, there were higher proportions of cases with hospitalizations, intensive care services, and mortality, compared to cases without pre-existing conditions (Table 1).For example, cases with CVD, compared to cases without CVD, were more likely to be hospitalized (17.5% vs 3.0%, respectively, p<0.01), to need ICU (0.9% vs, 0.2%, respectively, p <0.01), and to die from COVID-19 (6.9% vs 0.9%, respectively, p<0.01). Cases with diabetes, compared to cases without diabetes, were more likely to be hospitalized (21.4% vs 3.3%, respectively, p<0.01), more likely to need ICU (1.1% vs, 0.2%, respectively, p <0.01) and had higher mortality from COVID-19 (7.6% vs 1.0%, respectively, p<0.01). This pattern was observed across all pre-existing conditions (Table 2), and more so in cases with kidney disease, compared to cases without kidney disease, appeared to have very higher vulnerability, with high proportions hospitalized (44.7% vs 4.1%, respectively, p<0.01), in ICU (1.5% vs 0.2%, respectively, p<0.01) and high mortality (15.6% vs 1.3%, respectively, p<0.01)

**Table 1.**
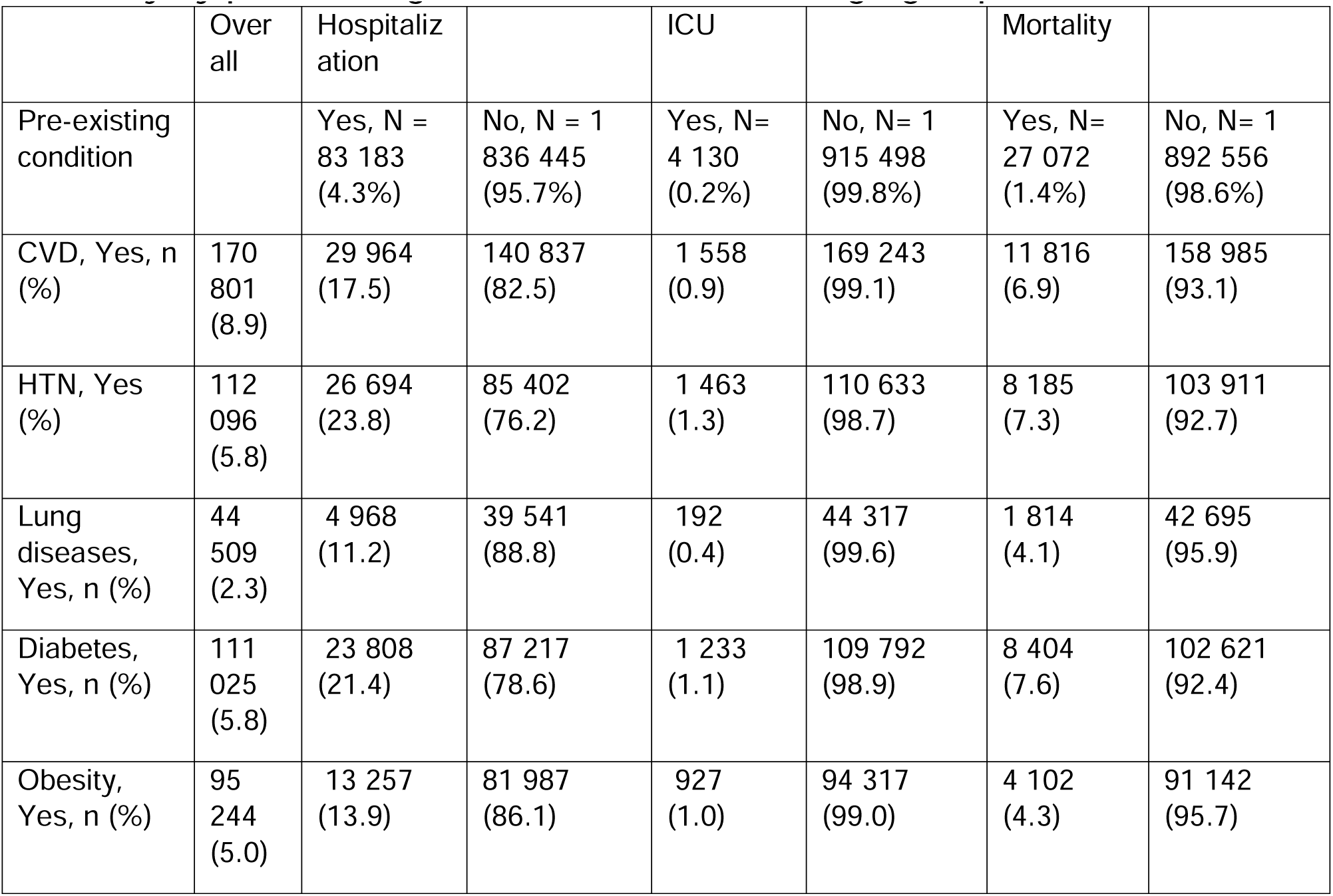
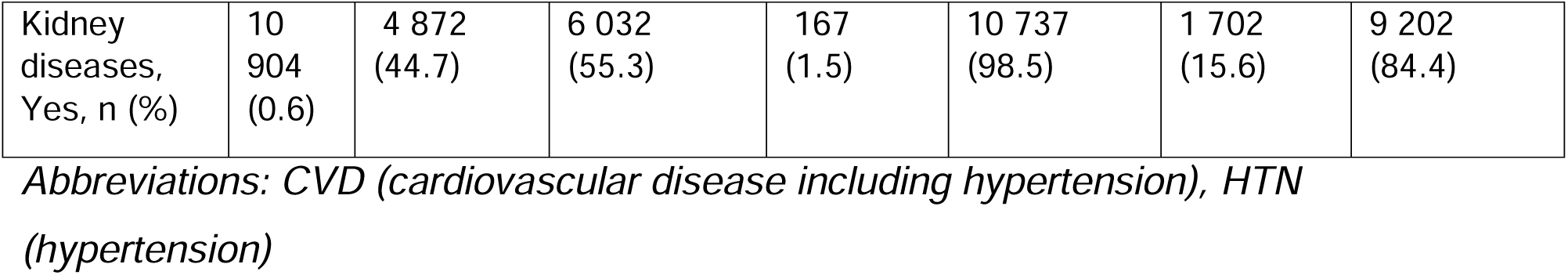
– Comparison of hospitalizations, intensive care services, and mortality by pre-existing condition status for all age groups.

|  | Over all | Hospitalization |  | ICU |  | Mortality |  |
| --- | --- | --- | --- | --- | --- | --- | --- |
| Pre-existing condition |  | Yes, N = 83 183 (4.3%) | No, N = 1 836 445 (95.7%) | Yes, N= 4 130 (0.2%) | No, N= 1 915 498 (99.8%) | Yes, N= 27 072 (1.4%) | No, N= 1 892 556 (98.6%) |
| CVD, Yes, n (%) | 170 801 (8.9) | 29 964 (17.5) | 140 837 (82.5) | 1 558 (0.9) | 169 243 (99.1) | 11 816 (6.9) | 158 985 (93.1) |
| HTN, Yes (%) | 112 096 (5.8) | 26 694 (23.8) | 85 402 (76.2) | 1 463 (1.3) | 110 633 (98.7) | 8 185 (7.3) | 103 911 (92.7) |
| Lung diseases, Yes, n (%) | 44 509 (2.3) | 4 968 (11.2) | 39 541 (88.8) | 192 (0.4) | 44 317 (99.6) | 1 814 (4.1) | 42 695 (95.9) |
| Diabetes, Yes, n (%) | 111 025 (5.8) | 23 808 (21.4) | 87 217 (78.6) | 1 233 (1.1) | 109 792 (98.9) | 8 404 (7.6) | 102 621 (92.4) |
| Obesity, Yes, n (%) | 95 244 (5.0) | 13 257 (13.9) | 81 987 (86.1) | 927 (1.0) | 94 317 (99.0) | 4 102 (4.3) | 91 142 (95.7) |
| Kidney diseases, Yes, n (%) | 10 904 (0.6) | 4 872 (44.7) | 6 032 (55.3) | 167 (1.5) | 10 737 (98.5) | 1 702 (15.6) | 9 202 (84.4) |

*Abbreviations: CVD (cardiovascular disease including hypertension), HTN (hypertension)*

**Table 2.** – Overall adjusted odds ratios of hospitalization, ICU admission, and mortality for each pre-existing conditions.

| Overall | Hospitalisation | ICU admission | Mortality |
| --- | --- | --- | --- |
|  | aOR (95% CI) | aOR (95% CI) | aOR (95% CI) |
| Cardiovascular diseases | 1.7 (1.7-1.7) | 1.4 (1.3-1.5) | 1.7 (1.6-1.7) |
| Lung Diseases | 1.9 (1.8-1.9) | 1.1 (0.9-1.3) | 1.6 (1.5-1.7) |
| Diabetes | 2.2 (2.1-2.2) | 1.7 (1.5-1.8) | 2.0 (1.9-2.0) |
| Kidney diseases | 5.5 (5.2-5.7) | 1.4 (1.2-1.7) | 2.7 (2.6-2.9) |
| Obesity | 1.7 (1.6-1.7) | 2.2 (2.1-2.4) | 1.9 (1.8-2.0) |
| Hypertension | 1.5 (1.4-1.5) | 1.3 (1.2-1.4) | 1.3 (1.3-1.4) |

### Comparison of proportions of hospitalizations, ICU and mortality by pre-existing condition presence in each age group

Overall, individuals with any of the pre-existing conditions were more likely to be hospitalized compared to those without pre-existing conditions [CVD (17.6% vs 2.0%, p<0.001), diabetes (21.5% vs 2.2%, p<0.001), lung diseases (11.3% vs 2.9%, p<0.001), kidney diseases (44.8% vs 2.8%, p<0.001), hypertension (23.9% vs 2.1%, p<0.001) and obesity (14.0% vs 2.6%, p<0.001)] (Supplementary Table 4). The difference in the proportions of hospitalized cases between those with and without pre-existing conditions was highest in the younger age groups and decreased in the older age groups (Supplementary Table 4).

Similarly, individuals with any of the pre-existing conditions were more likely to be in the ICU compared to those without pre-existing conditions [CVD (0.9% vs 0.1%, p<0.01), diabetes (1.1% vs 0.1%, p<0.001), lung diseases (0.4% vs 0.1%, p<0.001), kidney diseases (1.5% vs 0.1%, p<0.001), hypertension (1.3% vs 0.1%, p<0.001) and obesity (1.0% vs 0.1%, p<0.001)]. Again, the difference in the proportions of ICU admission between those with and without pre-existing conditions was highest in the younger age groups and lower in the older age groups (Supplementary Table 5).

Individuals with any of the pre-existing conditions were more likely to die from COVID-19 compared to those without pre-existing conditions [CVD (7.0% vs 0.6%, p<0.01), diabetes (7.6% vs 0.7%, p<0.001), lung diseases (4.2% vs 0.9%, p<0.001), kidney diseases (15.8% vs 0.9%, p<0.001), hypertension (7.3% vs 0.7%, p<0.001) and obesity (4.3% vs 0.9%, p<0.001)]. Overall, the difference of proportion of death between those with and without pre-existing conditions were highest in the younger age groups and lower in the older age groups (Supplementary Table 6).

### Association between pre-existing conditions and hospitalization, ICU admission and mortality from COVID-19

The overall adjusted odds of hospitalization for each pre-existing condition were; CVD (OR 1.7, 95%CI 1.7-1.7), hypertension (OR 1.5, 95%CI 1.4-1.5), diabetes (OR 2.2, 95%CI 2.1-2.2), obesity (OR 1.7, 95%CI 1.6-1.7), kidney disease (OR 5.5, 95%CI 5.2-5.7) and lung disease (OR 1.9, 95%CI 1.8-1.9) (Fig. 3) (Table 2 and Supplementary Table 7). Similarly, the overall adjusted odds of ICU admission for each pre-existing condition were; CVD (OR 2.1, 95%CI 1.8-2.4), hypertension (OR 1.3, 95%CI 1.2-1.4), diabetes (OR 1.7, 95%CI 1.5-1.8), obesity (OR 2.2, 95%%CI 2.1-2.4), kidney disease (OR 1.4, 95%CI 1.2-1.7) and lung disease (OR 1.1, 95%CI 0.9-1.3) (Fig. 3 and Supplementary Table 8). Finally, the overall adjusted odds of mortality for each pre-existing condition were; CVD (OR 1.7, 95%CI 1.6-1.7), hypertension (OR 1.3, 95%CI 1.3-1.4), diabetes (OR 2.0, 95%CI 1.9-2.0), obesity (OR 1.9, 95%CI 1.8-2.0), kidney disease (OR 2.7, 95%CI 2.6-2.9) and lung disease (OR 1.6, 95%CI 1.5-1.7) (Fig. 3 and Supplementary Table 9). The odds of each outcome were considerably larger in children and young adults with these preexisting conditions than for adults, especially for kidney disease, CVD, and diabetes. The odds ratio for hospitalization, ICU admission and mortality were very large for children and young adults (10-40 years) who had CVD, diabetes, hypertension, kidney disease (Fig 2 and Table 3). Also, the odds ratios for hospitalization, ICU admission and mortality were higher than for the other preexisting conditions up to the age group of 70-79 years (Fig 2).

**Fig. 2.**
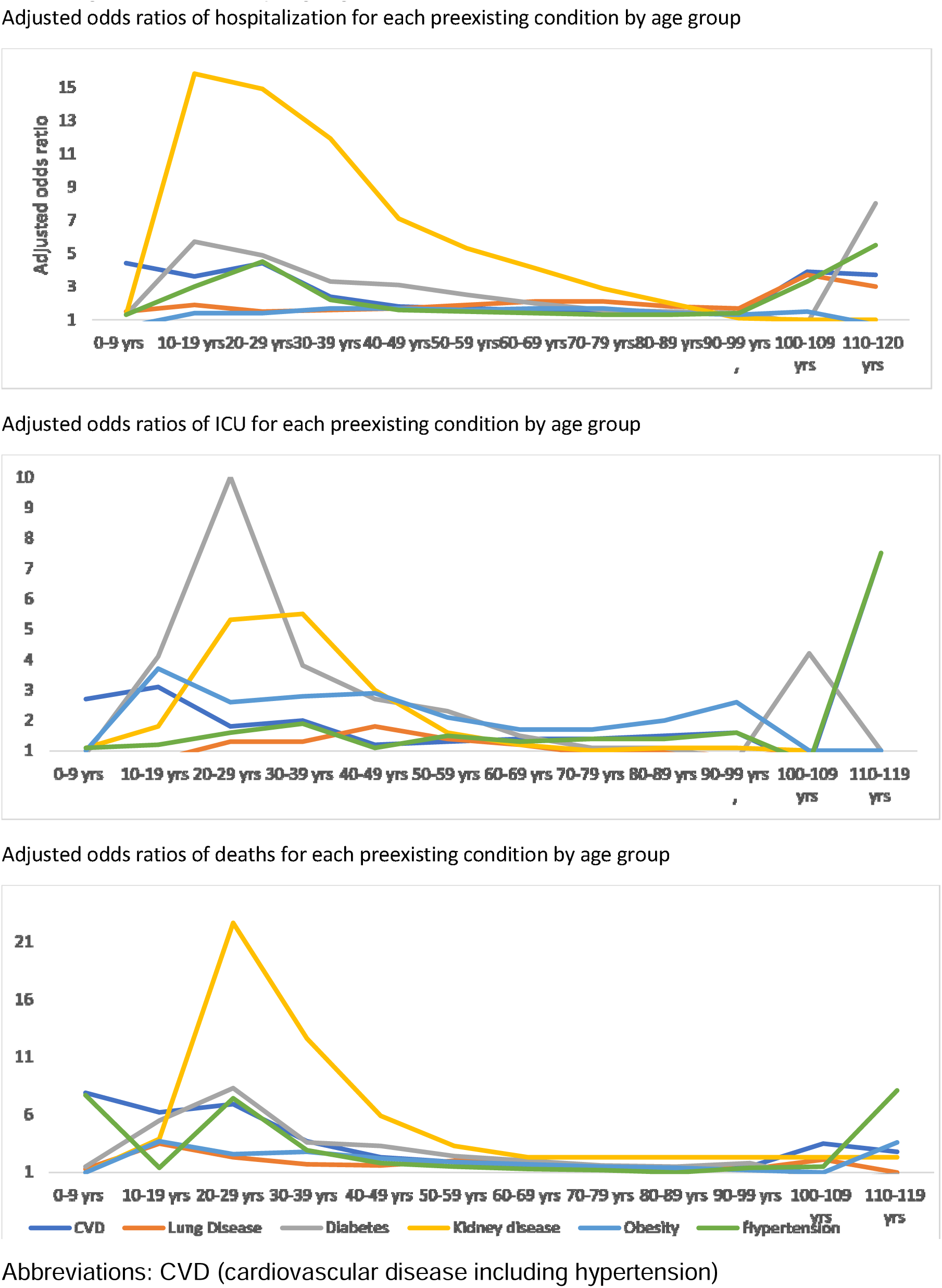
– Adjusted odds ratios of hospitalization, ICU and mortality for each pre-existing conditions by age group.

**Fig. 3.**
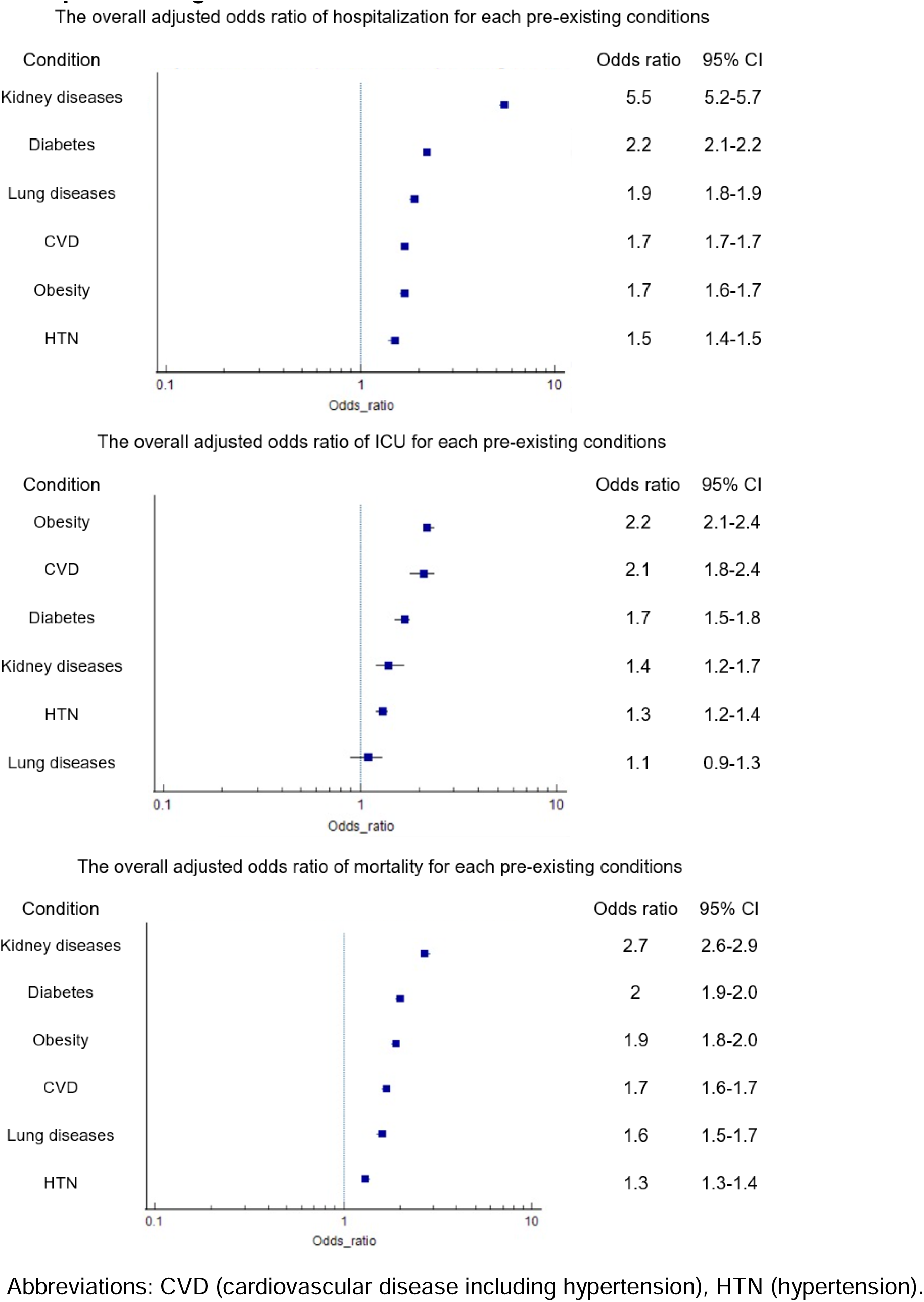
– The overall adjusted odds ratios of hospitalization, ICU and mortality for each pre-existing conditions.

## Discussion

In this cross-sectional analysis of the GH data repository, we confirmed that individuals with pre-existing CVD, diabetes, obesity, hypertension, kidney disease and lung diseases have higher odds of hospitalization, ICU admission and death from COVID-19 outcome compared to those with no pre-existing comorbidities. Kidney diseases were associated with the highest odds of mortality from COVID-19, followed by diabetes, obesity, CVD and lung diseases. Our findings were in alignment with other different studies (9, 20–24). meta-analysis reported that patient with renal diseases have overexpression of ACE2 receptors as well as immune system dysregulation that makes them more susceptible to severe COVID-19 infection and outcomes (25). The risk of preexisting conditions on different outcomes markedly varied between studies, including when compared to the findings of this study. The contrast in these findings might be a result of the contexts, measurements, and definitions of variables in individual primary studies as well as the issue of correlation or co-occurrence between pre-existing conditions. Since the early phases of the pandemic, in addition to socio-demographic factors, heart failure, kidney disease and obesity have been observed as strong risk factors for hospital admission and critical illness with COVID-19 (26, 27). What is clear and evidently demonstrated by this study and other similar studies is that approximately most of patients that experience worse COVID-19 outcomes have at least one pre-existing condition (28).

We found that children and young adults with pre-existing conditions have higher odds of severe COVID-19 outcomes. For example, children and young adults with kidney diseases had almost 20 folds increase in odds of COVID-19 mortality compared to those with no comorbidities. Of note, young adults aged 20-29 years had 10 times the odds of ICU admission due to COVID-19 infection compared to those with no comorbidities. Different studies have found that children tend to have a mild course of COVID-19 infection and a better prognosis than the older population. However, there is a paucity of research that assesses the effect of pre-existing conditions on COVID-19 prognosis among that age group. A meta-analysis reported that children with comorbidities are at a higher risk of COVID-19 mortality compared to healthy children. Despite reporting that obesity among children might be associated with worse COVID-19 outcomes, they did not determine the association between other comorbidities and COVID-19 outcomes among children. Therefore, using the Global health data repository, we were able to study the association between multiple well-defined comorbidities and clinical COVID-19 outcomes.

The findings of this study benefit from a large sample for comparison between patients. Nevertheless, the nature of being an observational study mean has several weaknesses which should be taken into consideration during interpretation. We used data from an international repository which was reported by governments. It is likely that some individuals with undiagnosed pre-existing conditions may have been missed. There is a possibility of ascertainment bias as data was collected mostly at the point of care, or at the hospital. It is likely not every patient who had severe COVID-19 visited healthcare facilities. There is a possibility that these patients might systematically or significantly differ from the sampled population.

This study used data from the Global Health Repository, a database of verifiable, international COVID-19 cases. With over 25 million cases as of March 20, 2020, the database serves as a large open-data resource for researchers investigating COVID-19. With a large proportion of cases coming from MICs and LICs, where cases were usually underreported, the repository now provides data that can influence the healthcare policy in countries with fewer resources. The use of Global Health Repository by researchers may be able to provide governments and healthcare sectors the needed data to target the high-risk populations, in addition to identifying weaknesses in the healthcare infrastructure across countries.

## Conclusion

This analysis of a global health repository confirms associations between pre-existing diseases and clinical outcomes of COVID-19. The odds of these outcomes are especially elevated in children and young adults with these preexisting conditions.

## Data Availability

All data produced in the present study are available upon reasonable request to the authors

