## Supplementary Table for "Association between pre-existing conditions and hospitalization, intensive care services and mortality from COVID-19 – a cross sectional analysis of an international global health data repository"

Supplementary Table 1 – Characteristics of cases in the global health data set as of 10th March 2021

|  | Hospitalization |  | ICU |  | Mortality |  | Total, N = 25 774 885 |
| --- | --- | --- | --- | --- | --- | --- | --- |
|  | Yes, N = 692 001 (2.7%) | No, N = 25 082 884 (97.3%) | Yes, N = 50 195 (0.2%) | No, N = 25 724 690 (99.8%) | Yes, N = 433 394 (1.7%) | No, N = 25 341 491 (98.3%) |  |
| Gender, Female, n (%) | 335 165 (2.8) | 11 853 326 (97.3) | 20 646 (0.2) | 12 167 845 (99.8) | 192 446 (1.6) | 11 996 045 (98.4) | 12 188 491 (47.3) |
| Gender, Male, n (%) | 354 059 (3.2) | 10 870 336 (96.9) | 29 433 (0.3) | 11 194 962 (99.7) | 240 130 (2.1) | 10 984 265 (97.9) | 11 224 395 (43.5) |
| Gender, Others, n (%) | 13 (4.3) | 287 (95.7) | 4 (1.3) | 296 (98.7) | 5 (1.7) | 295 (98.3) | 300 (0.0) |
| Gender, Missing, n(%) | 2 764 (0.1) | 2 358 935 (99.9) | 112 (0.0) | 2 361 587 (100.0) | 813 (0.0) | 2 360 886 (100.0) | 2 361 699 (9.2) |
| Year of diagnosis |  |  |  |  |  |  |  |
| 2020, n (%) | 557 643 (2.9) | 19 021 218 (97.2) | 43 021 (0.2) | 19 535 840 (99.8) | 366 883 (1.9) | 19 211 978 (98.1) | 19 578 861 (76.0) |
| 2021, n (%) | 134 358 (2.2) | 6 061 666 (97.8) | 7 174 (0.1) | 6 188 850 (99.9) | 66 511 (1.1) | 6 129 513 (98.9) | 6 196 024 (24.0) |
| Period of diagnosis |  |  |  |  |  |  |  |
| Jan-Mar 2020 | 27 372 (5.8) | 445 973 (94.2) | 3 264 (0.7) | 470 081 (99.3) | 11 807 (2.5) | 461 538 (97.5) | 473 345 (1.8) |
| Apr-Jun 2020 | 167 268 (4.7) | 3 399 956 (95.3) | 17 993 (0.5) | 3 549 231 (99.5) | 109 771 (3.1) | 3 457 453 (96.9) | 3 567 224 (13.8) |
| Jul- Sep 2020 | 122 355 (2.4) | 4 907 294 (97.6) | 9 271 (0.2) | 5 020 378 (99.8) | 92 153 (1.8) | 4 937 496 (98.2) | 5 029 649 (19.5) |
| Oct-Dec 2020 | 240 648 (2.3) | 10 267 995 (97.7) | 12 493 (0.1) | 10 496 150 (99.9) | 153 152 (1.5) | 10 355 491 (98.5) | 10508643 (40.8) |
| Jan-Mar 2021 | 134 358 (2.2) | 6 061 666 (97.8) | 7 174 (0.1) | 6 188 850 (99.9) | 66 511 (1.1) | 6 129 513 (98.9) | 6 196 024 (24.0) |
| Biggest case contributions by country |  |  |  |  |  |  |  |
| USA, n (%) | 587 195 (3.9) | 14 388 175 (96.1) | 43 152 (0.3) | 14 932 218 (99.7) | 259 753 (1.7) | 14 715 617 (98.3) | 14 975 370 (58.1) |
| Germany, n (%) | 0 | 2 448 424 (100.0) | 0 | 2 448 424 (100.0) | 65 191 (2.7) | 2 383 233 (97.3) | 2 448 424 (9.5) |
| Colombia, n (%) | 17 883 (0.8) | 2 211 189 (99.2) | 2 770 (0.1) | 2 226 302 (99.9) | 66 943 (3.0) | 2 162 129 (97.0) | 2 229 072 (8.6) |
| Brazil, n (%) | 6 286 (0.3) | 1 874 049 (99.7) | 61 (0.0) | 1 880 274 (100.0) | 9 016 (0.5) | 1 871 319 (99.5) | 1 880 335 (7.3) |

Supplementary Table 2 – Countries reporting pre-existing conditions

|  | pre-existing conditions |  |  |  |  |  | Outcome |  |  | Total |
| --- | --- | --- | --- | --- | --- | --- | --- | --- | --- | --- |
| Country | CVD | HTN | Diabetes | Lung diseases | Kidney diseases | Obesity | Hosp | ICU | Mortality |  |
|  | Yes | Yes | Yes | Yes | Yes | Yes | Yes | Yes | Yes |  |
| Brazil, n (%) | 55 782 (3.0) | 0 | 30 198 (1.6) | 19 569 (1.0) | 1 749 (0.1) | 4 092 (0.2) | 6 286 (0.3) | 61 (0.0) | 9 016 (0.5) | 1 880 335 |
| Canada, n (%) | 2 (0.0) | 2 (0.0) | 0 | 0 | 0 | 0 | 0 | 0 | 0 | 58 302 |
| Cuba, n (%) | 0 (0.00) | 0 | 3 (0.0) | 31 (0.1) | 0 | 0 | 0 | 0 | 0 | 22 183 |
| Gabon, n (%) | 1 (16.7) | 1 (16.7) | 1 (16.7) | 0 | 0 | 0 | 0 | 0 | 0 | 6 |
| India, n (%) | 1 (0.0) | 1 (0.0) | 1 (0.0) | 0 | 0 | 0 | 2 919 (14.4) | 0 | 33 (0.2) | 20 302 |
| Mexico, n (%) | 116 850 (13.1) | 112 382 (12.6) | 81 784 (9.2) | 25 434 (2.9) | 9 222 (1.0) | 91 230 (10.2) | 77 718 (8.7) | 4 083 (0.5) | 18 597 (2.1) | 893 738 |
| Moldova, n (%) | 1 (0.0) | 1 (0.0) | 1 (0.0) | 0 | 0 | 1 (0.0) | 0 | 0 | 0 | 11 067 |
| Nigeria, n (%) | 0 | 0 | 1 (0.0) | 0 | 0 | 0 | 0 | 0 | 0 | 6 403 |
| South Korea, n (%) | 0 | 0 | 0 | 2 (0.0) | 0 | 0 | 0 | 9 (0.1) | 36 (0.3) | 14 544 |
| Vietnam, n (%) | 1 (3.1) | 1 (3.1) | 1 (3.1) | 0 | 0 | 0 | 0 | 26 (81.3) | 0 | 32 |

“No” may contain missing

All percentages are rows

Abbreviations: CVD (cardiovascular disease including hypertension), HTN (hypertension), ICU (intensive care services)

Supplementary Table 3 - Characteristics and outcomes of cases from countries reporting pre-existing conditions (Brazil, Cuba, and Mexico)

|  | Overall | Hospitalization |  |  | ICU |  |  | Mortality |  |  |
| --- | --- | --- | --- | --- | --- | --- | --- | --- | --- | --- |
|  |  | Yes, N = 83 183 (4.3%) | No, N = 1 836 107 (95.7%) | P-value | Yes, N= 4 130 (0.2%) | No, N= 1 915 160 (99.8%) | P- value | Yes, N= 27 072 (1.4%) | No, N= 1 892 218 (98.6%) | P- value |
| Gender n (%) |  |  |  |  |  |  |  |  |  |  |
| Female | 1 009 988 (52.6) | 36 469 (3.6) | 973 519 (96.4) |  | 1 653 (0.2) | 1 008 335 (99.8) |  | 10 558 (1.1) | 999 430 (99.0) |  |
| Male | 909 302 (47.4) | 46 714 (5.1) | 862 588 (94.9) | <0.01 | 2 477 (0.3) | 906 825 (99.7) | <0.01 | 16 514 (1.8) | 892 788 (98.2) | <0.01 |
| Missing | 338 (0.0) |  |  |  |  |  |  |  |  |  |
| Age groups, n (%) |  |  |  |  |  |  |  |  |  |  |
| 0-9 yrs | 63 613 (3.3) | 4 350 (6.8) | 59 263 (93.2) |  | 252 (0.4) | 63 361 (99.6) |  | 158 (0.3) | 63 455 (99.8) |  |
| 10-19 yrs | 108 642 (5.7) | 1 957 (1.8) | 106 685 (98.2) |  | 76 (0.1) | 108 566 (99.9) |  | 83 (0.1) | 108 559 (99.9) |  |
| 20-29 yrs | 372 832 (19.4) | 4 279 (1.2) | 368 553 (98.9) |  | 165 (0.0) | 372 667 (100.0) |  | 375 (0.1) | 372 457 (99.9) |  |
| 30-39 yrs | 439 122 (22.9) | 6 930 (1.6) | 432 192 (98.4) |  | 300 (0.1) | 438 822 (99.9) |  | 1 038 (0.2) | 438 084 (99.8) |  |
| 40-49 yrs | 379 180 (19.8) | 11 064 (2.9) | 368 116 (97.1) |  | 505 (0.1) | 378 675 (99.9) |  | 2 626 (0.7) | 376 554 (99.3) |  |
| 50-59 yrs | 280 179 (14.6) | 15 707 (5.6) | 264 472 (94.4) |  | 807 (0.3) | 279 372 (99.7) |  | 4 831 (1.7) | 275 348 (98.3) |  |
| 60-69 yrs | 150 070 (7.8) | 17 863 (11.9) | 132 207 (88.1) |  | 941 (0.6) | 149 129 (99.4) |  | 6 492 (4.3) | 143 578 (95.7) |  |
| 70-79 yrs | 74 989 (3.9) | 13 611 (18.2) | 61 378 (81.9) |  | 725 (1.0) | 74 264 (99.0) |  | 6 135 (8.2) | 68 854 (91.8) |  |
| 80-89 yrs | 26 762 (1.4) | 6 445 (24.1) | 20 317 (75.9) |  | 305 (1.1) | 26 457 (98.9) |  | 3 236 (12.1) | 23 526 (87.9) |  |
| 90-99 yrs | 3 971 (0.2) | 923 (23.2) | 3 048 (76.8) |  | 47 (1.2) | 3 924 (98.8) |  | 491 (12.4) | 3 480 (87.6) |  |
| 100-109 yrs | 350 (0.0) | 36 (10.3) | 314 (89.7) |  | 5 (1.4) | 345 (98.6) |  | 25 (7.1) | 325 (92.9) |  |
| 110-120 yrs | 192 (0.0) | 18 (9.4) | 174 (90.6) | <0.01 | 2 (1.0) | 190 (99.0) | <0.01 | 14 (7.3) | 178 (92.7) | <0.01 |
| Country |  |  |  |  |  |  |  |  |  |  |
| Brazil, n (%) | 1 015 975 (52.9) | 5 729 (0.6) | 1 010 246 (99.4) |  | 61 (0.0) | 1 015 914 (100.0) |  | 5 569 (0.8) | 1 007 406 (99.2) |  |
| Mexico, n (%) | 893 167 (46.5) | 77 454 (8.7) | 815 713 (91.3) |  | 4 069 (0.5) | 889 098 (99.5) |  | 18 503 (2.1) | 874 664 (97.9) |  |
| Cuba, n (%) | 10 486 (0.5) | 0 | 10 486 (100.0) | <0.01 | 0 | 10 486 (100.0) | <0.01 | 0 | 10 486 (100.0) | <0.01 |
| Year of diagnosis |  |  |  |  |  |  |  |  |  |  |
| 2020, n (%) | 1 070 514 (55.8) | 26 892 (2.5) | 1 043 622 (97.5) |  | 1 666 (0.2) | 1 068 848 (99.8) |  | 15 301 (1.4) | 1 055 213 (98.6) |  |
| 2021, n (%) | 849 114 (44.2) | 56 291 (6.6) | 792 823 (93.4) | <0.01 | 2 464 (0.3) | 846 650 (99.7) | <0.01 | 11 771 (1.4) | 837 343 (98.6) | 0.01 |
| Period of diagnosis |  |  |  |  |  |  |  |  |  |  |
| Jan-Mar 2020 | 29 805 (1.6) | 6 232 (20.9) | 23 573 (79.1) |  | 86 (0.3) | 29 719 (99.7) |  | 838 (2.8) | 28 967 (97.2) |  |
| Apr-Jun 2020 | 336 763 (17.5) | 5 327 (1.6) | 331 436 (98.4) |  | 326 (0.1) | 336 437 (99.9) |  | 5 894 (1.8) | 330 869 (98.3) |  |
| Jul- Sep 2020 | 371 038 (19.3) | 5 927 (1.6) | 365 111 (98.4) |  | 400 (0.1) | 370 638 (99.9) |  | 3 958 (1.1) | 367 080 (98.9) |  |
| Oct-Dec 2020 | 332 908 (17.3) | 9 406 (2.8) | 323 502 (97.2) |  | 854 (0.3) | 332 054 (99.7) |  | 4 611 (1.4) | 328 297 (98.6) |  |
| Jan-Mar 2021 | 849 114 (44.2) | 56 291 (6.6) | 792 823 (93.4) | <0.01 | 2 464 (0.3) | 846 650 (99.7) | <0.01 | 11 771 (1.4) | 837 343 (98.6) | <0.01 |

Supplementary Table 4 - Proportion of cases with preexisting conditions hospitalized by age group

| Hospitalizations by age groups | Cardiovascular Disease |  |  | Diabetes |  |  | Lung Disease |  |  | Kidney Disease |  |  | Hypertension |  |  | Obesity |  |  |
| --- | --- | --- | --- | --- | --- | --- | --- | --- | --- | --- | --- | --- | --- | --- | --- | --- | --- | --- |
|  | Yes, n (%) | No, n (%) | P value | Yes, n (%) | No, n (% | P value | Yes, n (%) | No, n (%) | P value | Yes, n (%) | No, n (%) | P value | Yes, n (%) | No, n (%) | P value | Yes, n (%) | No, n (%) | P value |
| Overall, hospitalized | 30 403 (17.6) | 53 601 (2.0) |  | 24 019 (21.5) | 59 985 (2.2) |  | 5 090 (11.3) | 78 914 (2.9) |  | 4 915 (44.8) | 79 089 (2.8) |  | 26 836 (23.9) | 57 168 (2.1) |  | 13 301 (14.0) | 70 703 (2.6) |  |
| Overall, not hospitalized, | 142 229 (82.4) | 2 570 023 (98.0) | <0.01 | 87 966 (78.6) | 2 624 286 (97.8) | <0.01 | 39 944 (88.7) | 2 672 308 (97.1) | <0.01 | 6 056 (55.2) | 2 706 196 (97.2) | <0.01 | 85 546 (76.1) | 2 626 706 (97.9) | <0.01 | 82 021 (86.1) | 2 630 231 (97.4) | <0.01 |
| 0-9 yrs, hospitalized | 173 (33.1) | 4 177 (6.6) | <0.01 | 26 (15.6) | 4 324 (6.8) | <0.01 | 190 (13.1) | 4 160 (6.7) | <0.01 | 19 (24.1) | 4 331 (6.8) | <0.01 | 23 (28.8) | 4 327 (6.8) | <0.01 | 38 (14.1) | 4 312 (6.8) | <0.01 |
| 0-9 yrs, not hospitalized | 350 (66.9) | 58 913 (93.4) |  | 141 (84.4) | 59 122 (93.2) |  | 1 256 (86.9) | 58 007 (93.3) |  | 60 (76.0) | 59 203 (93.2) |  | 57 (71.3) | 59 206 (93.2) |  | 231 (85.9) | 59 032 (93.2) |  |
| 10-19 yrs, hospitalized | 66 (10.7) | 1 891 (1.8) | <0.01 | 67 (15.0 | 1 890 (1.8) | <0.01 | 114 (3.9) | 1 843 (1.7) | <0.01 | 71 (39.2) | 1 886 (1.7) | <0.01 | 38 (16.0) | 1 919 (1.8) | <0.01 | 104 (5.3) | 1 853 (1.7) | <0.01 |
| 10-19 yrs, not hospitalized | 549 (89.3) | 106 136 (98.3) |  | 379 (85.0) | 106 306 (98.3) |  | 2 823 (96.1) | 103 862 (98.3) |  | 110 (60.8) | 106 575 (98.3) |  | 199 (84.0) | 106 486 (98.2) |  | 1 853 (94.7) | 104 832 (98.3) |  |
| 20-29 yrs, hospitalized | 385 (8.22) | 3 894 (1.1) | <0.01 | 240 (11.0) | 4 039 (1.1) | <0.01 | 155 (2.0) | 4 124 (1.1) | <0.01 | 269 (30.9) | 4 010 (1.1) | <0.01 | 344 (11.7) | 3 935 (1.1) | <0.01 | 481 (3.4) | 3 798 (1.1) | <0.01 |
| 20-29 yrs, not hospitalized | 4 299 (91.8) | 364 254 (98.9) |  | 1 939 (89.0) | 366 614 (98.9) |  | 7 748 (98.0) | 360 805 (98.9) |  | 603 (69.2) | 367 950 (98.9) |  | 2 591 (88.3) | 365 962 (98.9) |  | 13 549 (96.6) | 355 004 (98.9) |  |
| 30-39 yrs, hospitalized | 884 (6.6) | 6 046 (1.4) | <0.01 | 695 (9.5) | 6 235 (1.4) | <0.01 | 241 (3.0) | 6 689 (1.6) | <0.01 | 402 (32.4) | 6 528 (1.5) | <0.01 | 773 (9.4) | 6 157 (1.4) | <0.01 | 1 252 (5.7) | 5 678 (1.4) | <0.01 |
| 30-39 yrs, not hospitalized | 12 434 (93.4) | 419 758 (98.6) |  | 6 635 (90.5) | 425 557 (98.6) |  | 7 847 (97.0) | 424 345 (98.5) |  | 838 (67.6) | 431 354 (98.5) |  | 7 468 (90.6) | 424 724 (98.6) |  | 20 800 (94.3) | 411 392 (98.6) |  |
| 40-49 yrs, hospitalized | 2 585 (8.7) | 8 479 (2.4) | <0.01 | 2 543 (13.1) | 8 521 (2.4) | <0.01 | 413 (5.5) | 10 651 (2.9) | <0.01 | 550 (35.7) | 10 514 (2.8) | <0.01 | 2 315 (11.7) | 8 749 (2.4) | <0.01 | 2 335 (10.2) | 8 729 (2.5) | <0.01 |
| 40-49 yrs, not hospitalized | 27 250 (91.3) | 340 866 (97.6) |  | 16 820 (86.9) | 351 296 (97.6) |  | 7 086 (94.5) | 361 030 (97.1) |  | 990 (64.3) | 367 126 (97.2) |  | 17 454 (88.3) | 350 662 (97.6) |  | 20 616 (89.8) | 347 500 (97.6) |  |
| 50-59 yrs, hospitalized | 5 747 (12.9) | 9 960 (4.2) | <0.01 | 5 284 (17.0) | 10 423 (4.2) | <0.01 | 679 (11.2) | 15 028 (5.5) | <0.01 | 1 000 (44.4) | 14 707 (5.3) | <0.01 | 5 176 (17.2) | 10 531 (4.2) | <0.01 | 3 130 (17.1) | 12 577 (4.8) | <0.01 |
| 50-59 yrs, not hospitalized | 38 723 (87.1) | 225 749 (95.8) |  | 25 827 (83.0) | 238 645 (95.8) |  | 5 410 (88.9) | 259 062 (94.5) |  | 1 250 (55.6) | 263 222 (94.7) |  | 24 972 (82.8) | 239 500 (95.8) |  | 15 187 (82.9) | 249 285 (95.2) |  |
| 60-69 yrs, hospitalized | 8 524 (22.0) | 9 339 (8.4) | <0.01 | 7 207 (25.4) | 10 656 (8.8) | <0.01 | 1 084 (23.8) | 16 779 (11.5) | <0.01 | 1 308 (56.3) | 16 555 (11.2) | <0.01 | 7 633 (28.6) | 10 230 (8.3) | <0.01 | 3 176 (32.4) | 14 687 (10.5) | <0.01 |
| 60-69 yrs, not hospitalized | 30 307 (78.1) | 101 900 (91.6) |  | 21 148 (74.6) | 111 059 (91.3) |  | 3 463 (76.2) | 128 744 (88.5) |  | 1 013 (43.6) | 131 194 (88.8) |  | 19 017 (71.4) | 113 190 (91.7) |  | 6 615 (67.6) | 125 592 (89.5) |  |
| 70-79 yrs, hospitalized | 7 420 (30.3) | 6 191 (12.3) | <0.01 | 5 383 (33.1) | 8 228 (14.0) | <0.01 | 1 176 (34.5) | 12 435 (17.4) | <0.01 | 843 (58.8) | 12 768 (17.4) | <0.01 | 6 564 (40.1) | 7 047 (12.0) | <0.01 | 1 988 (46.9) | 11 623 (16.4) | <0.01 |
| 70-79 yrs, not hospitalized | 17 058 (69.7) | 44 320 (87.7) |  | 10 875 (66.9) | 50 503 (86.0) |  | 2 229 (65.5) | 59 149 (82.6) |  | 590 (41.2) | 60 788 (82.6) |  | 9 810 (59.9) | 51 568 (88.0) |  | 2 252 (53.1) | 59 126 (83.6) |  |
| 80-89 yrs, hospitalized | 3 676 (38.2) | 2 769 (16.2) | <0.01 | 2 151 (40.6) | 4 294 (20.0) | <0.01 | 785 (46.0) | 5 660 (22.6) | <0.01 | 369 (59.0) | 6 076 (23.3) | <0.01 | 3 363 (49.6) | 3 082 (15.4) | <0.01 | 689 (53.0) | 5 756 (22.6) | <0.01 |
| 80-89 yrs, not hospitalized | 5 937 (61.8) | 14 380 (83.9) |  | 3 151 (59.4) | 17 166 (80.0) |  | 922 (54.0) | 19 395 (77.4) |  | 256 (41.0) | 20 061 (76.8) |  | 3 420 (50.4) | 16 897 (84.6) |  | 610 (47.0) | 19 707 (77.4) |  |
| 90-99 yrs, hospitalized | 483 (40.8) | 440 (15.8) | <0.01 | 201 (42.1) | 722 (20.7) | <0.01 | 122 (44.0) | 801 (21.7) | <0.01 | 37 (53.6) | 886 (22.7) | <0.01 | 446 (53.4) | 477 (15.2) | <0.01 | 61 (55.0) | 862 (22.3) | <0.01 |
| 90-99 yrs, not hospitalized | 702 (59.2) | 2 346 (84.2) |  | 277 (58.0) | 2 771 (79.3) |  | 155 (56.0) | 1 893 (78.3) |  | 32 (46.4) | 3 016 (77.3) |  | 390 (46.7) | 2 658 (84.8) |  | 50 (45.1) | 2 998 (77.7) |  |
| 100-109 yrs, hospitalized | 15 (25.4) | 21 (7.2) | <0.01 | 6 (26.1) | 30 (9.2) | 0.02 | 6 (31.6) | 30 (9.1) | 0.01 | 4 (100.0) | 32 (9.3) | <0.01 | 13 (43.3) | 23 (7.2) | <0.01 | 2 (33.3) | 34 (9.9) | 0.12 |
| 100-109 yrs, not hospitalized | 44 (74.6) | 270 (92.8) |  | 17 (73.9) | 297 (90.8) |  | 13 (68.4) | 301 (90.9) |  | 0 (0.0) | 314 (90.8) |  | 17 (56.7) | 297 (92.8) |  | 4 (66.7) | 310 (90.1) |  |
| 110-120 yrs, hospitalized | 6 (22.2) | 12 (7.3) | 0.03 | 5 (38.5) | 13 (7.3) | <0.01 | 3 (30.0) | 15 (8.2) | 0.06 | NA | 18 | NA | 6 (46.2) | 12 (6.7) | <0.01 | 1 (20.0) | 17 (9.1) | 0.39 |
| 110-120 yrs, not hospitalized | 21 (77.8) | 153 (92.7) |  | 8 (61.5) | 166 (92.7) |  | 7 (70.0) | 167 (91.8) |  | NA | 174 |  | 7 (53.9) | 167 (93.3) |  | 4 (80.0) | 170 (90.9) |  |

Supplementary Table 5 - Proportion of cases with preexisting conditions in ICU by age group

| ICU by age groups | Cardiovascular disease |  |  | Diabetes |  |  | Lung Disease |  |  | Kidney Disease |  |  | Hypertension |  |  | Obesity |  |  |
| --- | --- | --- | --- | --- | --- | --- | --- | --- | --- | --- | --- | --- | --- | --- | --- | --- | --- | --- |
|  | Yes, n (%) | No, n (%) | P value | Yes, n (%) | No, n (%) | P value | Yes, n (%) | No, n (%) | P value | Yes, n (%) | No, n (%) | P value | Yes, n (%) | No, n (%) | P value | Yes, n (%) | No, n (%) | P value |
| Overall, ICU | 1 567 (0.9) | 2 577 (0.1) |  | 1 236 (1.1) | 2 908 (0.1) |  | 194 (0.4) | 3 950 (0.1) |  | 168 (1.5) | 3 976 (0.1) |  | 1 472 (1.3) | 2 672 (0.1) |  | 928 (1.0) | 3 216 (0.1) |  |
| Overall, No ICU | 171 065 (99.1) | 2 621 047 (99.9) | <0.01 | 110 749 (98.9) | 2 681 363 (99.9) | <0.01 | 44 840 (99.6) | 2 747 272 (99.9) | <0.01 | 10 803 (98.5) | 2 781 309 (99.9) | <0.01 | 110 910 (98.7) | 2 681 202 (99.9) | <0.01 | 94 394 (99.0) | 2 697 718 (99.9) | <0.01 |
| 0-9 yrs ICU | 10 (1.9) | 242 (0.4) | <0.01 | 1 (0.6) | 251 (0.4) | 0.49 | 2 (0.1) | 250 (0.4) | 0.11 | 1 (1.3) | 251 (0.4) | 0.27 | 1 (1.3) | 251 (0.4) | 0.27 | 0 | 252 (0.4) | 0.63 |
| 0-9 yrs, No ICU | 513 (98.1) | 62 848 (99.6) |  | 166 (99.4) | 63 195 (99.6) |  | 1 444 (99.9) | 61 917 (99.6) |  | 78 (98.7) | 63 283 (99.6) |  | 79 (98.8) | 63 282 (99.6) |  | 269 (100.0) | 63 092 (99.6) |  |
| 10-19 yrs, ICU | 3 (0.5) | 73 (0.1) | 0.01 | 3 (0.7) | 73 (0.1) | <0.01 | 2 (0.1) | 74 (0.1) | 1.00 | 1 (0.6) | 75 (0.1) | 0.12 | 1 (0.4) | 75 (0.1) | 0.15 | 10 (0.5) | 66 (0.1) | <0.01 |
| 10-19 yrs, No ICU | 612 (99.5) | 107 954 (99.9) |  | 443 (99.3) | 108 123 (99.9) |  | 2 935 (99.9) | 105 631 (99.9) |  | 180 (99.5) | 108 386 (99.9) |  | 236 (99.6) | 108 330 (99.9) |  | 1 947 (99.5) | 106 619 (99.9) |  |
| 20-29 yrs, ICU | 12 (0.3) | 153 (0.0) | <0.01 | 18 (0.8) | 147 (0.0) | <0.01 | 6 (0.1) | 159 (0.0) | 0.17 | 7 (0.8) | 158 (0.0) | <0.01 | 10 (0.3) | 155 (0.0) | <0.01 | 32 (0.2) | 133 (0.0) | <0.01 |
| 20-29 yrs, No ICU | 4 672 (99.7) | 367 996 (100.0) |  | 2 161 (99.2) | 370 506 (100.0) |  | 7 897 (99.9) | 364 770 (100.0) |  | 865 (99.2) | 371 802 (100.0) |  | 2 925 (99.7) | 369 742 (100.0) |  | 13 998 (99.8) | 358 669 (100.0) |  |
| 30-39 yrs, ICU | 43 (0.3) | 257 (0.1) | <0.01 | 41 (0.6) | 259 (0.1) | <0.01 | 9 (0.1) | 291 (0.1) | 0.14 | 16 (1.3) | 284 (0.1) | <0.01 | 40 (0.5) | 260 (0.1) | <0.01 | 86 (0.4) | 214 (0.1) | <0.01 |
| 30-39 yrs, No ICU | 13 275 (99.7) | 425 547 (99.9) |  | 7 289 (99.4) | 431 533 (99.9) |  | 8 079 (99.9) | 430 743 (99.9) |  | 1 224 (98.7) | 437 598 (99.9) |  | 8 201 (99.5) | 430 621 (99.9) |  | 21 966 (99.6) | 416 856 (100.0) |  |
| 40-49 yrs, ICU | 110 (0.4) | 395 (0.1) | <0.01 | 122 (0.6) | 383 (0.1) | <0.01 | 22 (0.3) | 483 (0.1) | <0.01 | 19 (1.2) | 486 (0.1) | <0.01 | 101 (0.5) | 404 (0.1) | <0.01 | 162 (0.7) | 343 (0.1) | <0.01 |
| 40-49 yrs, No ICU | 29 725 (99.6) | 348 950 (99.9) |  | 19 241 (99.4) | 359 434 (99.9) |  | 7 477 (99.7) | 371 198 (99.9) |  | 1 521 (98.8) | 377 154 (99.9) |  | 19 688 (99.5) | 359 007 (99.9) |  | 22 789 (99.3) | 355 886 (99.9) |  |
| 50-59 yrs, ICU | 284 (0.6) | 523 (0.2) | <0.01 | 289 (0.9) | 518 (0.2) | <0.01 | 30 (0.5) | 777 (0.3) | <0.01 | 31 (1.4) | 776 (0.3) | <0.01 | 273 (0.9) | 534 (0.2) | <0.01 | 207 (1.1) | 600 (0.2) | <0.01 |
| 50-59 yrs, No ICU | 44 186 (99.4) | 235 186 (99.8) |  | 30 822 (99.1) | 248 550 (99.8) |  | 6 059 (99.5) | 273 313 (99.7) |  | 2 219 (98.6) | 277 153 (99.7) |  | 29 875 (99.1) | 249 497 (99.8) |  | 18 110 (98.9) | 261 262 (99.8) |  |
| 60-69 yrs, ICU | 469 (1.2) | 472 (0.4) | <0.01 | 379 (1.3) | 562 (0.5) | <0.01 | 45 (1.0) | 896 (0.6) | <0.01 | 44 (1.9) | 897 (0.6) | <0.01 | 443 (1.7) | 498 (0.4) | <0.01 | 232 (2.4) | 709 (0.5) | <0.01 |
| 60-69 yrs, No ICU | 38 362 (98.8) | 110 767 (99.6) |  | 27 976 (98.7) | 121 153 (99.5) |  | 4 502 (99.0) | 144 627 (99.4) |  | 2 277 (98.1) | 146 852 (99.4) |  | 26 207 (98.3) | 122 922 (99.6) |  | 9 559 (97.6) | 139 570 (99.5) |  |
| 70-79 yrs, ICU | 409 (1.7) | 316 (0.6) | <0.01 | 269 (1.7) | 456 (0.8) | <0.01 | 42 (1.2) | 683 (1.0) | 0.10 | 31 (2.2) | 694 (0.9) | <0.01 | 391 (2.4) | 334 (0.6) | <0.01 | 140 (3.3) | 585 (0.8) | <0.01 |
| 70-79 yrs, No ICU | 24 069 (98.3) | 50 195 (99.4) |  | 15 989 (98.4) | 58 275 (99.2) |  | 3 363 (98.8) | 70 901 (99.1) |  | 1 402 (97.8) | 72 862 (99.1) |  | 15 983 (97.6) | 58 281 (99.4) |  | 4 100 (96.7) | 70 164 (99.2) |  |
| 80-89 yrs, ICU | 189 (2.0) | 116 (0.7) | <0.01 | 102 (1.9) | 203 (1.0) | <0.01 | 30 (1.8) | 275 (1.1) | 0.01 | 15 (2.4) | 290 (1.1) | <0.01 | 176 (2.6) | 129 (0.7) | <0.01 | 52 (4.0) | 253 (1.0) | <0.01 |
| 80-89 yrs, No ICU | 9 424 (98.0) | 17 033 (99.3) |  | 5 200 (98.1) | 21 257 (99.1) |  | 1 677 (98.2) | 24 780 (98.9) |  | 610 (97.6) | 25 847 (98.9) |  | 6 607 (97.4) | 19 850 (99.4) |  | 1 247 (96.0) | 25 210 (99.0) |  |
| 90-99 yrs, ICU | 27 (2.3) | 20 (0.7) | <0.01 | 8 (1.7) | 39 (1.1) | 0.29 | 4 (1.4) | 43 (1.2) | 0.57 | 2 (2.9) | 45 (1.2) | 0.20 | 25 (3.0) | 22 (0.7) | <0.01 | 6 (5.4) | 41 (1.1) | <0.01 |
| 90-99 yrs, No ICU | 1 158 (97.7) | 2 766 (99.3) |  | 470 (98.3) | 3 454 (98.9) |  | 273 (98.6) | 3 651 (98.8) |  | 67 (97.1) | 3 857 (98.9) |  | 811 (97.0) | 3 113 (99.3) |  | 105 (94.6) | 3 819 (98.9) |  |
| 100-109 yrs, ICU | 1 (1.7) | 4 (1.4) | 1.00 | 1 (4.4) | 4 (1.2) | 0.29 | 0 | 5 (1.5) | 1.00 | 0 | 5 (1.5) | 1.00 | 1 (3.3) | 4 (1.3) | 0.36 | 0 | 5 (1.5) | 1.00 |
| 100-109 yrs, No ICU | 58 (98.3) | 287 (98.6) |  | 22 (95.7) | 323 (98.8) |  | 19 (100.0) | 326 (98.5) |  | 4 (100.0) | 341 (98.6) |  | 29 (96.7) | 316 (98.8) |  | 6 (100.0) | 339 (98.6) |  |
| 110-120 yrs, ICU | 1 (3.7) | 1(0.6) | 0.26 | 0 | 2 (1.2) | 1.00 | 0 | 2 (1.1) | 1.00 | NA | 2 (1.0) | NA | 1 (7.7) | 1 (0.6) | 0.13 | 0 | 2 (1.1) | 1.00 |
| 110-120 yrs, No ICU | 26 (96.3) | 164 (99.4) |  | 13 (100.0) | 177 (98.9) |  | 10 (100.0) | 180 (98.9) |  | NA | 190 (99.0) |  | 12 (92.3) | 178 (99.4) |  | 5 (100.0) | 185 (98.9) |  |

Supplementary Table 6 - Proportion of deaths with preexisting conditions by age group

| Mortality by age groups | Cardiovascular Disease |  |  | Diabetes |  |  | Lung Disease |  |  | Kidney Disease |  |  | Hypertension |  |  | Obesity |  |  |
| --- | --- | --- | --- | --- | --- | --- | --- | --- | --- | --- | --- | --- | --- | --- | --- | --- | --- | --- |
|  | Yes, n (%) | No, n (%) | P value | Yes, n (%) | No, n (%) | P value | Yes, n (%) | No, n (%) | P value | Yes, n (%) | No, n (%) | P value | Yes, n (%) | No, n (%) | P value | Yes, n (%) | No, n (%) | P value |
| Overall, deaths | 12 074 (7.0) | 15 539 (0.6) |  | 8 548 (7.6) | 19 065 (0.7) |  | 1 888 (4.2) | 25 725 (0.9) |  | 1 733 (15.8) | 25 880 (0.9) |  | 8 238 (7.3) | 19 375 (0.7) |  | 4 126 (4.3) | 23 487 (0.9) |  |
| Overall, alive | 160 558 (93.0) | 2 608 085 (99.4) | <0.01 | 103 437 (92.4) | 2 665 206 (99.3) | <0.01 | 43 146 (95.8) | 2 725 497 (99.1) | <0.01 | 9 238 (84.2) | 2 759 405 (99.1) | <0.01 | 104 144 (92.7) | 2 664 499 (99.3) | <0.01 | 91 196 (95.7) | 2 677 447 (99.1) | <0.01 |
| 0-9 yrs deaths | 15 (2.9) | 143 (0.2) | <0.01 | 2 (1.2) | 156 (1.3) | 0.07 | 6 (0.4) | 152 (0.2) | 0.18 | 0 | 158 (0.3) | 1.00 | 4 (5.0) | 154 (0.2) | <0.01 | 2 (0.7) | 156 (0.3) | 0.14 |
| 0-9 yrs alive | 508 (97.1) | 62 947 (99.8) |  | 165 (98.8) | 63 290 (99.8) |  | 1 440 (99.6) | 62 015 (99.8) |  | 79 (100.0) | 63 376 (99.8) |  | 76 (95.0) | 63 379 (99.8) |  | 267 (99.3) | 63 188 (99.8) |  |
| 10-19 yrs, deaths | 5 (0.8) | 78 (0.1) | <0.01 | 4 (0.9) | 79 (0.1) | <0.01 | 8 (0.3) | 75 (0.1) | <0.01 | 2 (1.1) | 81 (0.1) | 0.01 | 1 (0.4) | 82 (0.1) | 0.17 | 7 (0.4) | 76 (0.1) | <0.01 |
| 10-19 yrs, alive | 610 (99.2) | 107 949 (99.9) |  | 442 (99.1) | 108 117 (99.9) |  | 2 929 (99.7) | 105 630 (99.9) |  | 179 (98.9) | 108 380 (99.9) |  | 236 (99.6) | 108 323 (99.9) |  | 1 950 (99.6) | 106 609 (99.9) |  |
| 20-29 yrs, deaths | 60 (1.3) | 315 (0.1) | <0.01 | 43 (2.0) | 332 (0.1) | <0.01 | 20 (0.3) | 355 (0.1) | <0.01 | 50 (5.7) | 325 (0.1) | <0.01 | 51 (1.7) | 324 (0.1) | <0.01 | 59 (0.4) | 316 (0.1) | <0.01 |
| 20-29 yrs, alive | 4 624 (98.7) | 367 833 (99.9) |  | 2 136 (98.0) | 370 321 (99.9) |  | 7 883 (99.8) | 364 574 (99.9) |  | 822 (94.3) | 371 635 (99.9) |  | 2 884 (98.3) | 369 573 (99.9) |  | 13 971 (99.6) | 358 486 (99.9) |  |
| 30-39 yrs, deaths | 194 (1.5) | 844 (0.2) | <0.01 | 127 (1.7) | 911 (0.2) | <0.01 | 35 (0.4) | 1 003 (0.2) | <0.01 | 94 (7.6) | 944 (0.2) | <0.01 | 140 (1.7) | 898 (0.2) | <0.01 | 231 (1.1) | 807 (0.2) | <0.01 |
| 30-39 yrs, alive | 13 124 (98.5) | 424 960 (99.8) |  | 7 203 (98.3) | 430 881 (99.8) |  | 8 053 (99.6) | 430 031 (99.8) |  | 1 146 (92.4) | 436 938 (99.8) |  | 8 101 (98.3) | 429 983 (99.8) |  | 21 821 (99.0) | 416 263 (99.8) |  |
| 40-49 yrs, deaths | 736 (2.5) | 1 1890 (0.5) | <0.01 | 662 (3.4) | 1 964 (0.6) | <0.01 | 90 (1.2) | 2 536 (0.7) | <0.01 | 170 (11.0) | 2 456 (0.7) | <0.01 | 564 (2.9) | 2 062 (0.6) | <0.01 | 594 (2.6) | 2 032 (0.6) | <0.01 |
| 40-49 yrs, alive | 29 099 (97.5) | 347 455 (99.5) |  | 18 701 (96.6) | 357 853 (99.5) |  | 7 409 (98.8) | 369 145 (99.3) |  | 1 370 (89.0) | 375 184 (99.4) |  | 19 205 (97.2) | 357 349 (99.4) |  | 22 357 (97.4) | 354 197 (99.4) |  |
| 50-59 yrs, deaths | 1 838 (4.1) | 2 993 (1.3) | <0.01 | 1 603 (5.2) | 3 228 (1.3) | <0.01 | 223 (3.7) | 4 608 (1.7) | <0.01 | 300 (13.3) | 4 531 (1.6) | <0.01 | 1 413 (4.7) | 3 418 (1.4) | <0.01 | 981 (5.4) | 3 850 (1.5) | <0.01 |
| 50-59 yrs, alive | 42 632 (95.9) | 232 716 (98.7) |  | 29 508 (94.9) | 245 840 (98.7) |  | 5 866 (96.3) | 269 482 (98.3) |  | 1 950 (86.7) | 273 398 (98.4) |  | 28 735 (95.3) | 246 613 (98.6) |  | 17 336 (94.6) | 258 012 (98.5) |  |
| 60-69 yrs, deaths | 3 021 (7.8) | 3 471 (3.1) | <0.01 | 2 593 (9.1) | 3 899 (3.2) |  | 402 (8.8) | 6 090 (4.2) | <0.01 | 428 (18.4) | 6 064 (4.1) | <0.01 | 2 420 (9.1) | 4 072 (3.3) | <0.01 | 1 070 (10.9) | 5 422 (3.9) | <0.01 |
| 60-69 yrs, alive | 35 810 (92.2) | 107 768 (96.9) |  | 25 762 (90.9) | 117 816 (96.8) |  | 4 145 (91.2) | 139 433 (95.8) |  | 1 893 (81.6) | 141 685 (95.9) |  | 24 230 (90.9) | 119 348 (96.7) |  | 8 721 (89.1) | 134 857 (96.1) |  |
| 70-79 yrs, deaths | 3 090 (12.6) | 3 045 (6.0) | <0.01 | 2 290 (14.1) | 3 845 (6.6) | <0.01 | 461 (13.5) | 5 674 (7.9) | <0.01 | 335 (23.4) | 5 800 (7.9) | <0.01 | 2 324 (14.2) | 3 811 (6.5) | <0.01 | 750 (17.7) | 5 385 (7.6) | <0.01 |
| 70-79 years, alive | 21 388 (87.4) | 47 466 (94.0) |  | 13 968 (85.9) | 54 886 (93.5) |  | 2 944 (86.5) | 65 910 (92.1) |  | 1 098 (76.6) | 67 756 (92.1) |  | 14 050 (85.8) | 54 804 (93.5) |  | 3 490 (82.3) | 65 364 (92.4) |  |
| 80-89 yrs, deaths | 1 548 (16.1) | 1 688 (9.8) | <0.01 | 968 (18.3) | 2 268 (10.6) | <0.01 | 303 (17.8) | 2 933 (11.7) | <0.01 | 148 (23.7) | 3 088 (11.8) | <0.01 | 1 094 (16.1) | 2 142 (10.7) | <0.01 | 265 (20.4) | 2 971 (11.7) | <0.01 |
| 80-89 yrs, alive | 8 065 (83.9) | 15 461 (90.2) |  | 4 334 (81.7) | 19 192 (89.4) |  | 1 404 (82.3) | 22 122 (88.3) |  | 477 (76.3) | 23 049 (88.2) |  | 5 689 (83.9) | 17 837 (89.3) |  | 1 034 (79.6) | 22 492 (88.3) |  |
| 90-99 yrs, deaths | 222 (18.7) | 269 (9.7) | <0.01 | 110 (23.0) | 381 (10.9) | <0.01 | 49 (17.7) | 442 (12.0) | 0.01 | 16 (23.2) | 475 (12.2) | 0.01 | 170 (20.3) | 321 (10.2) | <0.01 | 24 (21.6) | 467 (12.1) | <0.01 |
| 90-99 yrs, alive | 963 (81.3) | 2 517 (90.3) |  | 368 (77.0) | 3 112 (89.1) |  | 228 (82.3) | 3 252 (88.0) |  | 53 (76.8) | 3 427 (87.8) |  | 666 (79.7) | 2 814 (89.8) |  | 87 (78.4) | 3 393 (87.9) |  |
| 100-109 yrs, deaths | 8 (13.6) | 17 (5.8) | 0.05 | 1 (4.4) | 24 (7.3) | 1.00 | 2 (10.5) | 23 (7.0) | 0.64 | 0 | 25 (7.2) | 1.00 | 1 (3.3) | 24 (7.5) | 0.71 | 0 | 25 (7.3) | 1.00 |
| 100-109 yrs, alive | 51 (86.4) | 274 (94.2) |  | 22 (95.7) | 303 (92.7) |  | 17 (89.5) | 308 (93.1) |  | 4 (100.0) | 321 (92.8) |  | 29 (96.7) | 296 (92.5) |  | 6 (100.0) | 319 (92.7) |  |
| 110-120 yrs, deaths | 4 (14.8) | 10 (6.1) | 0.12 | 1 (7.7) | 13 (7.3) | 1.00 | 0 | 14 (7.7) | 1.00 | NA | 14 (7.3) | NA | 3 (23.1) | 11 (6.2) | 0.06 | 1 (20.0) | 13 (7.0) | 0.32 |
| 110-120 yrs, alive | 23 (85.2) | 155 (93.9) |  | 12 (92.3) | 166 (92.7) |  | 10 (100.0) | 168 (92.3) |  | NA | 178 (92.7) |  | 10 (76.9) | 168 (93.9) |  | 4 (80.0) | 174 (93.1) |  |

Supplementary Table 7- Unadjusted and adjusted odds ratios for hospitalization for each preexisting condition by age group–multivariable logistic regression

| Age groups | Cardiovascular diseases |  | Lung Diseases |  | Diabetes |  | Kidney diseases |  | Obesity |  | Hypertension |  |
| --- | --- | --- | --- | --- | --- | --- | --- | --- | --- | --- | --- | --- |
|  | OR (95% CI) | aOR (95% CI) | OR (95% CI) | aOR (95% CI) | OR (95% CI) | aOR (95% CI) | OR (95% CI) | aOR (95% CI) | OR (95% CI) | aOR (95% CI) | OR (95% CI) | aOR (95% CI) |
| Overall | 6.8 (6.7-6.9) | 1.7 (1.7-1.7) | 2.9 (2.8-3.0) | 1.9 (1.8-1.9) | 8.0 (7.9-8.2) | 2.2 (2.1-2.2) | 18.9 (18.2-19.6) | 5.5 (5.2-5.7) | 4.1 (4.0-4.1) | 1.7 (1.6-1.7) | 9.7 (9.5-9.8) | 1.5 (1.4-1.5) |
| 0-9 yrs | 7.0 (5.8-8.4) | 4.4 (3.5-5.5) | 2.1 (1.8-2.5) | 1.5 (1.3-1.8) | 2.5 (1.7-3.8) | 1.3 (0.8-2.2) | 4.3 (2.6-7.3) | 1.3 (0.7-2.3) | 2.3 (1.6-3.2) | 0.6 (0.4-0.8) | 5.5 (3.4-9.0) | 1.3 (0.8-2.2) |
| 10-19 yrs | 6.7 (5.2-8.7) | 3.6 (2.7-4.8) | 2.3 (1.9-2.8) | 1.9 (1.5-2.3) | 9.9 (7.6-12.9) | 5.7 (4.3-7.7) | 36.5 (27.0-49.3) | 15.8 (11.4-22.1) | 3.2 (2.6-3.9) | 1.4 (1.1-1.7) | 10.6 (7.5-15.0) | 3.0 (2.0-4.4) |
| 20-29 yrs | 8.4 (7.5-9.3) | 4.4 (3.9-5.0) | 1.8 (1.5-2.1) | 1.5 (1.2-1.7) | 11.2 (9.8-12.9) | 4.9 (4.2-5.7) | 40.9 (35.3-47.4) | 14.9 (12.6-17.6) | 3.3 (3.0-3.7) | 1.4 (1.3-1.6) | 12.3 (11.0-13.9) | 4.5 (4.0-5.1) |
| 30-39 yrs | 4.9 (4.6-5.3) | 2.4 (2.2-2.6) | 1.9 (1.7-2.2) | 1.6 (1.4-1.8) | 7.1 (6.6-7.8) | 3.3 (3.0-3.6) | 31.7 (28.1-35.8) | 11.9 (10.4-13.7) | 4.4 (4.1-4.6) | 1.7 (1.6-1.8) | 7.1 (6.6-7.7) | 2.2 (2.1-2.4) |
| 40-49 yrs | 3.8 (3.6-4.0) | 1.8 (1.7-1.9) | 2.0 (1.8-2.2) | 1.7 (1.5-1.9) | 6.2 (5.9-6.5) | 3.1 (3.0-3.3) | 19.4 (17.4-21.6) | 7.1 (6.3-8.0) | 4.5 (4.3-4.7) | 1.7 (1.7-1.8) | 5.3 (5.1-5.6) | 1.6 (1.5-1.7) |
| 50-59 yrs | 3.4 (3.3-3.5) | 1.7 (1.6-1.8) | 2.2 (2.0-2.3) | 1.9 (1.7-2.0) | 4.7 (4.5-4.9) | 2.5 (2.4-2.5) | 14.3 (13.2-15.6) | 5.3 (4.8-5.8) | 4.1 (3.9-4.3) | 1.6 (1.6-1.7) | 4.7 (4.5-4.9) | 1.5 (1.5-1.6) |
| 60-69 yrs | 3.1 (3.0-3.2) | 1.6 (1.5-1.6) | 2.4 (2.2-2.6) | 2.1 (1.9-2.3) | 3.6 (3.4-3.7) | 2.0 (1.9-2.1) | 10.2 (9.4-11.1) | 4.1 (3.7-4.5) | 4.1 (3.9-4.3) | 1.7 (1.6-1.8) | 4.4 (4.3-4.6) | 1.4 (1.3-1.4) |
| 70-79 yrs | 3.1 (3.0-3.2) | 1.6 (1.5-1.6) | 2.5 (2.3-2.7) | 2.1 (2.0-2.3) | 3.0 (2.9-3.2) | 1.6 (1.6-1.7) | 6.8 (6.1-7.6) | 2.9 (2.5-3.2) | 4.5 (4.2-4.8) | 1.7 (1.6-1.8) | 4.9 (4.7-5.1) | 1.3 (1.2-1.3) |
| 80-89 yrs | 3.2 (3 .0-3.4) | 1.5 (1.4-1.6) | 2.9 (2.6-3.2) | 1.8 (1.6-2.1) | 2.7 (2.6-2.9) | 1.5 (1.4-1.6) | 4.8 (4.0-5.6) | 2.0 (1.7-2.4) | 3.9 (3.5-4.3) | 1.4 (1.2-1.6) | 5.4 (5.1-5.7) | 1.3 (1.2-1.4) |
| 90-99 yrs | 3.7 (3.1-4.3) | 1.4 (1.2-1.7) | 2.8 (2.2-3.6) | 1.7 (1.2-2.3) | 2.8 (2.3-3.4) | 1.4 (1.1-1.8) | 3.9 (2.4-6.4) | 1.1 (0.7-1.9) | 4.2 (2.9-6.2) | 1.3 (0.9-2.0) | 6.4 (5.4-7.5) | 1.4 (1.1-1.7) |
| 100-109 yrs | 4.4 (2.1-9.1) | 3.9 (1.5-9.7) | 4.6 (1.6-13.1) | 3.7 (1.1-12.5) | 3.5 (1.3-9.5) | 0.8 (0.2-3.1) | 1 (NA) | 1 (NA) | 4.6 (0.8-25.8) | 1.5 (0.2-10.5) | 9.9 (4.3-22.8) | 3.3 (1.2-8.9) |
| 110-120 yrs | 3.6 (1.2-10.7) | 3.7 (1.0-12.8) | 4.8 (1.1-20.4) | 3.0 (0.7-14.1) | 8.0 (2.3-27.9) | 8.0 (1.8-35.4) | 1 (NA) | 1 (NA) | 2.5 (0.3-23.7) | 0.7 (0.1-8.9) | 11.9 (3.5-41.1) | 5.5 (1.4-21.1) |

CVD adjusted for gender, country, diabetes and obesity. Lung diseases adjusted for gender, country and obesity. Diabetes adjusted for gender, country, CVD and obesity. Kidney diseases adjusted for gender, country, CVD, diabetes and obesity. Obesity adjusted forgender, country, CVD and diabetes. HTN adjusted for gender, country, diabetes and obesity.

Supplementary Table 8- Unadjusted and adjusted odds ratios for ICU for each preexisting condition by age group–multivariable logistic regression

| Age groups | Cardiovascular diseases |  | Lung Diseases |  | Diabetes |  | Kidney Diseases |  | Obesity |  | Hypertension |  |
| --- | --- | --- | --- | --- | --- | --- | --- | --- | --- | --- | --- | --- |
|  | OR (95% CI) | aOR (95% CI) | OR (95% CI) | aOR (95% CI) | OR (95% CI) | aOR (95% CI) | OR (95% CI) | aOR (95% CI) | OR (95% CI) | aOR (95% CI) | OR (95% CI) | aOR (95% CI) |
| Overall | 6.3 (5.9-6.7) | 1.4 (1.3-1.5) | 2.1 (1.8-2.4) | 1.1 (0.9-1.3) | 7.0 (6.5-7.5) | 1.7 (1.5-1.8) | 7.5 (6.4-8.7) | 1.4 (1.2-1.7) | 5.6 (5.2-6.0) | 2.2 (2.1-2.4) | 8.9 (8.4-9.5) | 1.3 (1.2-1.4) |
| 0-9 yrs | 5.1 (2.7-9.6) | 2.7 (1.4-5.1) | 0.3 (0.1-1.4) | 0.2 (0.1-1.0) | 1.5 (0.2-10.9) | 0.9 (0.1-6.7) | 3.2 (0.4-23.3) | 1.1 (0.1-8.1) | NA | NA | 3.2 (0.4-23.0) | 1.1 (0.1-7.9) |
| 10-19 yrs | 7.2 (2.3-23.1) | 3.1 (0.9-11.0) | 1.0 (0.2-4.0) | 0.7 (0.2-3.0) | 10.0 (3.1-31.9) | 4.1 (1.1-14.8) | 8.0 (1.1-58.1) | 1.8 (0.2-15.3) | 8.3 (4.3-16.2) | 3.7 (1.9-7.4) | 6.1 (0.8-44.2) | 1.2 (0.1-10.1) |
| 20-29 yrs | 6.2 (3.4-11.1) | 1.8 (1.0-3.5) | 1.7 (0.8-3.9) | 1.3 (0.6-3.0) | 21.0 (12.8-34.3) | 10.0 (5.8-17.3) | 19.0 (8.9-40.7) | 5.3 (2.2-12.7) | 6.2 (4.2-9.1) | 2.6 (1.7-3.8) | 8.2 (4.3-15.5) | 1.6 (0.8-3.2) |
| 30-39 yrs | 5.4 (3.9-7.4) | 2.0 (1.4-2.8) | 1.6 (0.8-3.2) | 1.3 (0.7-2.5) | 9.4 (6.7-13.0) | 3.8 (2.7-5.4) | 20.1 (12.1-33.4) | 5.5 (3.1-9.6) | 7.6 (5.9-9.8) | 2.8 (2.2-3.6) | 8.1 (5.8-11.3) | 1.9 (1.3-2.7) |
| 40-49 yrs | 3.3 (2.6-4.0) | 1.2 (1.0-1.6) | 2.3 (1.5-3.5) | 1.8 (1.2-2.8) | 6.0 (4.9-7.3) | 2.7 (2.2-3.4) | 9.7 (6.1-15.4) | 3.0 (1.8-4.8) | 7.4 (6.1-8.9) | 2.9 (2.4-3.5) | 4.6 (3.7-5.7) | 1.1 (0.9-1.4) |
| 50-59 yrs | 2.9 (2.5-3.3) | 1.3 (1.1-1.5) | 1.7 (1.2-2.5) | 1.4 (1.0-2.0) | 4.5 (3.9-5.2) | 2.3 (2.0-2.7) | 5.0 (3.5-7.2) | 1.6 (1.1-2.3) | 5.0 (4.2-5.8) | 2.1 (1.7-2.4) | 4.7 (4.5-4.9) | 1.5 (1.5-1.6) |
| 60-69 yrs | 2.9 (2.5-3.3) | 1.4 (1.2-1.7) | 1.6 (1.2-2.2) | 1.2 (0.9-1.6) | 2.9 (2.6-3.3) | 1.5 (1.3-1.7) | 3.2 (2.3-4.3) | 1.2 (0.9-1.6) | 4.1 (3.9-4.3) | 1.7 (1.6-1.8) | 4.2 (3.7-4.7) | 1.3 (1.2-1.5) |
| 70-79 yrs | 2.7 (2.3-3.1) | 1.4 (1.2-1.6) | 1.3 (0.9-1.8) | 0.9 (0.7-1.3) | 2.2 (1.8-2.5) | 1.1 (0.9-1.3) | 2.3 (1.6-3.3) | 1.0 (0.7-1.5) | 4.5 (4.2-4.8) | 1.7 (1.6-1.8) | 4.3 (3.7-4.9) | 1.4 (1.2-1.6) |
| 80-89 yrs | 2.9 (2.3-3.7) | 1.5 (1.2-1.9) | 1.6 (1.1-2.4) | 1.0 (0.7-1.4) | 2.1 (1.6-2.6) | 1.1 (0.9-1.4) | 2.2 (1.3-3.7) | 1.1 (0.6-1.8) | 4.2 (3.1-5.6) | 2.0 (1.5-2.7) | 4.1 (3.3-5.2) | 1.4 (1.1-1.7) |
| 90-99 yrs | 3.2 (1.8-5.8) | 1.6 (0.9-3.0) | 1.2 (0.4-3.5) | 0.8 (0.3-2.1) | 1.5 (0.7-3.2) | 0.7 (0.3-1.6) | 2.6 (0.6-10.8) | 1.1 (0.2-4.6) | 5.3 (2.2-12.8) | 2.6 (1.1-6.5) | 4.4 (2.4-7.8) | 1.6 (0.9-2.9) |
| 100-109 yrs | 1.2 (0.1-11.3) | 0.5 (0.0-8.1) | 1 (NA) | 1 (NA) | 3.7 (0.4-34.3) | 4.2 (0.3-70.2) | 1 (NA) | 1 (NA) | 1 (NA) | 1 (NA) | 2.7 (0.3-25.2) | 0.6 (0.0-11.1) |
| 110-120 yrs | 6.3 (0.4-104.0) | 7.5 (0.4-132.7) | 1 (NA) | 1 (NA) | 1 (NA) | 1 (NA) | 1 (NA) | 1 (NA) | 1 (NA) | 1 (NA) | 14.8 (0.9-252.1) | 7.5 (0.4-132.7) |

CVD adjusted for gender, country, diabetes and obesity. Lung diseases adjusted for gender, country and obesity. Diabetes adjusted for gender, country, CVD and obesity. Kidney diseases adjusted for gender, country, CVD, diabetes and obesity. Obesity adjusted for gender, country, CVD and diabetes. HTN adjusted for gender, country, diabetes and obesity.

Table 10 - Unadjusted and adjusted odds ratios for mortality for each preexisting condition by age group–multivariable logistic regression

| Age groups | Cardiovascular diseases |  | Lung Diseases |  | Diabetes |  | Kidney Diseases |  | Obesity |  | Hypertension |  |
| --- | --- | --- | --- | --- | --- | --- | --- | --- | --- | --- | --- | --- |
|  | OR (95% CI) | aOR (95% CI) | OR (95% CI) | aOR (95% CI) | OR (95% CI) | aOR (95% CI) | OR (95% CI) | aOR (95% CI) | OR (95% CI) | aOR (95% CI) | OR (95% CI) | aOR (95% CI) |
| Overall | 8.4 (8.2-8.7) | 1.7 (1.6-1.7) | 3.1 (3.0-3.3) | 1.6 (1.5-1.7) | 7.9 (7.6-8.1) | 2.0 (1.9-2.0) | 13.7 (13.0-14.5) | 2.7 (2.6-2.9) | 3.5 (3.4-3.7) | 1.9 (1.8-2.0) | 7.5 (7.3-7.7) | 1.3 (1.3-1.4) |
| 0-9 yrs, n (%) | 13.0 (7.6-22.3) | 7.9 (4.5-13.8) | 1.7 (0.8-3.9) | 1.3 (0.6-3.0) | 4.9 (1.2-20.0) | 1.5 (0.3-6.6) | 1 (NA) | 1 (NA) | 3.0 (0.7-12.3) | 1.0 (0.2-4.2) | 21.7 (7.8-59.9) | 7.7 (2.5-24.0) |
| 10-19 yrs, n (%) | 11.3 (4.6-28.1) | 6.2 (2.2-17.6) | 3.8 (1.9-8.0) | 3.5 (1.7-7.4) | 12.4 (4.5-34.0) | 5.5 (1.7-17.6) | 15.0 (2.6-61.2) | 3.9 (0.7-20.6) | 5.0 (2.3-10.9) | 3.7 (1.6-8.7) | 5.6 (0.8-40.4) | 1.4 (0.2-12.3) |
| 20-29 yrs, n (%) | 15.2 (11.5-20.0) | 6.9 (4.9-9.6) | 2.6 (1.7-4.1) | 2.3 (1.5-3.6) | 22.5 (16.3-30.9) | 8.3 (5.7-12.1) | 69.6 (51.2-94.4) | 22.6 (15.1-33.9) | 4.8 (3.6-6.3) | 2.6 (1.9-3.6) | 20.2 (15.0-27.2) | 7.4 (5.1-10.7) |
| 30-39 yrs, n (%) | 7.4 (6.4-8.7) | 3.7 (3.1-4.4) | 1.9 (1.3-2.6) | 1.7 (1.2-2.3) | 8.3 (6.9-10.1) | 3.6 (2.9-4.4) | 38.0 (30.5-47.3) | 12.6 (9.7-16.3) | 5.5 (4.7-6.3) | 2.8 (2.4-3.3) | 8.3 (6.9-9.9) | 2.9 (2.3-3.5) |
| 40-49 yrs, n (%) | 4.6 (4.3-5.1) | 2.3 (2.1-2.6) | 1.8 (1.4-2.2) | 1.6 (1.3-2.0) | 6.4 (5.9-7.1) | 3.3 (3.0-3.6) | 19.0 (16.1-22.3) | 5.9 (4.9-7.1) | 4.6 (4.2-5.1) | 2.2 (2.0-2.4) | 5.1 (4.6-5.6) | 1.8 (1.6-2.0) |
| 50-59 yrs, n (%) | 3.4 (3.2-3.6) | 1.9 (1.8-2.1) | 2.2 (1.9-2.5) | 2.0 (1.7-2.3) | 4.1 (3.9-4.4) | 2.4 (2.2-2.5) | 9.3 (8.2-10.5) | 3.3 (2.9-3.8) | 3.8 (3.5-4.1) | 1.9 (1.8-2.1) | 3.5 (3.3-3.8) | 1.5 (1.4-1.6) |
| 60-69 yrs, n (%) | 2.6 (2.5-2.8) | 1.6 (1.5-1.7) | 2.2 (2.0-2.5) | 1.9 (1.7-2.1) | 3.0 (2.9-3.2) | 2.0 (1.8-2.1) | 5.3 (4.7-5.9) | 2.3 (2.0-2.6) | 3.1 (2.8-3.3) | 1.7 (1.5-1.8) | 2.9 (2.8-3.1) | 1.3 (1.2-1.4) |
| 70-79 yrs, n (%) | 2.3 (2.1-2.4) | 1.5 (1.4-1.6) | 1.8 (1.6-2.0) | 1.6 (1.4-1.8) | 2.3 (2.2-2.5) | 1.6 (1.5-1.7) | 3.6 (3.1-4.0) | 1.9 (1.6-2.1) | 2.6 (2.4-2.8) | 1.5 (1.4-1.7) | 2.4 (2.3-2.5) | 1.2 (1.1-1.3) |
| 80-89 yrs, n (%) | 1.8 (1.6-1.9) | 1.4 (1.2-1.5) | 1.6 (1.4-1.9) | 1.4 (1.2-1.6) | 1.9 (1.7-2.1) | 1.5 (1.4-1.7) | 2.3 (1.9-2.8) | 1.5 (1.3-1.9) | 1.9 (1.7-2.2) | 1.4 (1.2-1.6) | 1.6 (1.5-1.7) | 1.0 (0.9-1.1) |
| 90-99 yrs , n (%) | 2.2 (1.8-2.6) | 1.5 (1.2-1.9) | 1.6 (1.1-2.2) | 1.3 (0.9-1.8) | 2.4 (1.9-3.1) | 1.8 (1.4-2.3) | 2.2 (1.2-3.8) | 1.1 (0.6-2.0) | 2.0 (1.3-3.2) | 1.2 (0.8-2.0) | 2.2 (1.8-2.7) | 1.4 (1.1-1.8) |
| 100-109 yrs, n (%) | 2.5 (1.0-6.2) | 3.5 (1.3-9.1) | 1.6 (0.3-7.2) | 2.1 (0.4-10.0) | 0.6 (0.1-4.4) | 0.4 (0.1-3.9) | 1 (NA) | 1 (NA) | 1 (NA) | 1 (NA) | 0.4 (0.1-3.3) | 1.5 (0.1-16.6) |
| 110-119 yrs, n (%) | 2.7 (0.8-9.3) | 2.8 (0.8-9.9) | 1 (NA) | 1 (NA) | 1.1 (0.1-8.8) | 0.8 (0.1-7.1) | 1 (NA) | 1 (NA) | 3.3 (0.3-32.2) | 3.6 (0.3-39.2) | 4.6 (1.1-19.1) | 8.1 (1.4-46.6) |

CVD adjusted for gender, country, diabetes and obesity. Lung diseases adjusted for gender, country and obesity. Diabetes adjusted for gender, country, CVD and obesity. Kidney diseases adjusted for gender, country, CVD, diabetes and obesity. Obesity adjusted for gender, country, CVD and diabetes. HTN adjusted for gender, country, diabetes and obesity.
